# Anticipating and assessing adverse and other unintended consequences of public health interventions: the (CONSEQUENT) framework

**DOI:** 10.1101/2023.02.03.23285408

**Authors:** Jan M Stratil, Renke L Biallas, Ani Movsisyan, Kathryn Oliver, Eva A Rehfuess

## Abstract

Despite the best intentions public health interventions (PHIs) can have adverse and other unintended consequences (AUCs). AUCs are rarely systematically examined when developing, evaluating or implementing PHIs. We used a structured, multi-pronged and evidence-based approach to develop a framework to support researchers and decision-makers in conceptualising and categorising AUCs of PHIs.

We employed the ‘best-fit’ framework synthesis approach. We designed the a-priori framework using elements of the WHO-INTEGRATE framework and the Behaviour Change Wheel. Next, we conducted a qualitative systematic review of theoretical and conceptual publications on the AUCs of PHIs in the databases Medline and Embase as well as through grey literature searches. Based on these findings, we iteratively revised and advanced the a-priori framework based on thematic analysis of the identified research. To validate and further refine the framework, we coded four systematic reviews on AUCs of distinct PHIs against it.

The CONSEQUENT framework includes two components: the first focuses on AUCs and serves to categorise them; the second component highlights the mechanisms through which AUCs may arise. The first component comprises eight domains of consequences – health, health system, human and fundamental rights, acceptability and adherence, equality and equity, social and institutional, economic and resource, and ecological.

The CONSEQUENT framework is intended to facilitate conceptualisation and categorising of AUCs of PHIs during their development, evaluation and implementation to support evidence-informed decision-making.

## 2 Research Manuscript section

### 2.1 Introduction

Promoting and improving the physical and mental health of populations is the central goal of public health (PH) interventions all over the globe. However, despite the best intentions, these interventions can have adverse effects, such as effects in the opposite direction of that intended or expected (paradoxical effects) or effects on unrelated outcomes (unintended externalities) (1). For example, providing pre-exposure prophylaxis against HIV may lead to an increase in risky sexual behaviour and in sexually transmitted infections other than HIV (2). The drilling of groundwater wells, which successfully reduced diarrheal disease mortality due to polluted surface water, has exposed an estimated 40 million Bangladeshis to harmful concentrations of arsenic contained in the groundwater (3). It has also been shown how obesity-focused PH interventions have led to an increase in stigmatisation and social exclusion of those living with obesity (4, 5).

To truly promote public health, it is essential not only to evaluate intended beneficial outcomes of PH interventions, but also to anticipate and assess their possible adverse and other unintended consequences (AUCs). In contrast to the well-established assessment of adverse drug reactions (ADRs), particular challenges arise in the assessment of AUCs of PH interventions: while ADRs primarily result directly from the drugs themselves and affect those taking the drugs, PH interventions often function as “events in systems”(6), where effects of the intervention arise as a result of the interaction between the intervention and the social, economic, or political context in which it is implemented (6-8). Individuals and populations not targeted by the intervention may even be those (most severely) affected by AUCs (9, 10). While ADRs are mostly health-related, PH interventions usually have social, economic, ecological, or political ramifications (e.g., large scale usage of DDT in malaria prevention leading to adverse effects on the ecosystem) (11-14). Furthermore, consideration of an unintended effect of an intervention as adverse, beneficial, or neutral is not always clear, as it depends on the perspective of the observer, as well as underlying sociocultural norms in different regions and populations; both perspectives and the sociocultural norms underlying them may change over time. For example, whether increased meat consumption is considered an adverse effect beyond (human) health is likely to depend on whether the evaluating person works in the meat industry or is an animal rights activist, whether the assessment takes place in Argentina or Nepal, and on the time of evaluation, such as in the 1980s compared to the 2020s.

Anticipating and understanding AUCs should be a priority for those deciding on or implementing PH interventions – as there are moral, ethical, political, and practical reasons for avoiding health and societal harms (15-17). However, these are often not thoroughly examined in public health research, practice, and policy, especially AUCs not directly related to health (18-20). While unintended consequences of social action have been discussed in the broader scientific literature (21-29), they constitute a largely neglected topic in empirical public health research (30, 31), except for specific areas, such as cancer screening (32) or illicit drug use (33).

In recent years, public health researchers have begun to identify and describe harms and to suggest typologies or classifications of harms (30, 31). However, these have primarily focused on health rather than broader societal consequences and/or have not been developed in a systematic manner (15, 34). Important questions remain on how to identify the unintended and potentially harmful effects of PH interventions (20), how best to evaluate them (19, 26), and how to incorporate the consideration of harms into the process of evidence-informed decision-making (14, 20, 35, 36). Being able to identify PH interventions and policies with substantive harmful effects and to subsequently revise, adapt or de-implement these interventions is essential for programme implementers, service providers, and policymakers.

The primary objective of the research project was to develop a framework which supports public health researchers, practitioners and decision-makers in conceptualising and categorising foreseeable AUCs of PH interventions (the *consequences* component of framework). The secondary objective was to map and conceptualise the mechanisms through which AUCs may arise (as a supplementary *mechanisms* component of framework).

In this context, we refer to conceptualising as the use of the framework as a tool to support stakeholders in systematically reflecting on (potential) AUCs of PH interventions when developing, evaluating, or implementing PH interventions. In the meantime, we refer to categorising as the use of the framework as a classification system of AUCs of PH interventions, for example when creating evidence (gap) maps.

### 2.2 Materials and Methods

#### 2.2.1 Overview of framework development process

We developed the framework using a “best-fit” framework synthesis approach (38, 39), starting with an a priori framework based on limited evidence, followed by iterative revisions of the framework considering further evidence. We used key elements from the WHO-INTEGRATE framework (36) and the Behaviour Change Wheel (40) to create an a priori framework of AUCs and the possible mechanisms leading to these (38, 39). We then advanced and refined the framework based on theoretical and conceptual papers describing frameworks or systems of AUCs of PH interventions and/or their mechanisms, as well as empirical research on the AUCs of PH interventions implemented in policy and practice. These papers were identified using systematic searches in health databases and reference searches (Supplementary Files 1-3). Thematic analysis was used to identify new themes and topics and thereby to revise the framework. In the final step, the findings in systematic reviews of the AUCs of four specific PH interventions were coded against the empirically advanced framework components (41-44), which were conducted by or in cooperation with the members of the research team. This served to validate the framework using examples from practice. The framework revisions across all steps were guided by discussions within the study team. The entire framework development process is visualised in Figure 1. The terminology used is defined in Box 1. We used the Standards for Reporting Qualitative Research (SRQR) reporting guideline (37).

**Figure 1.**
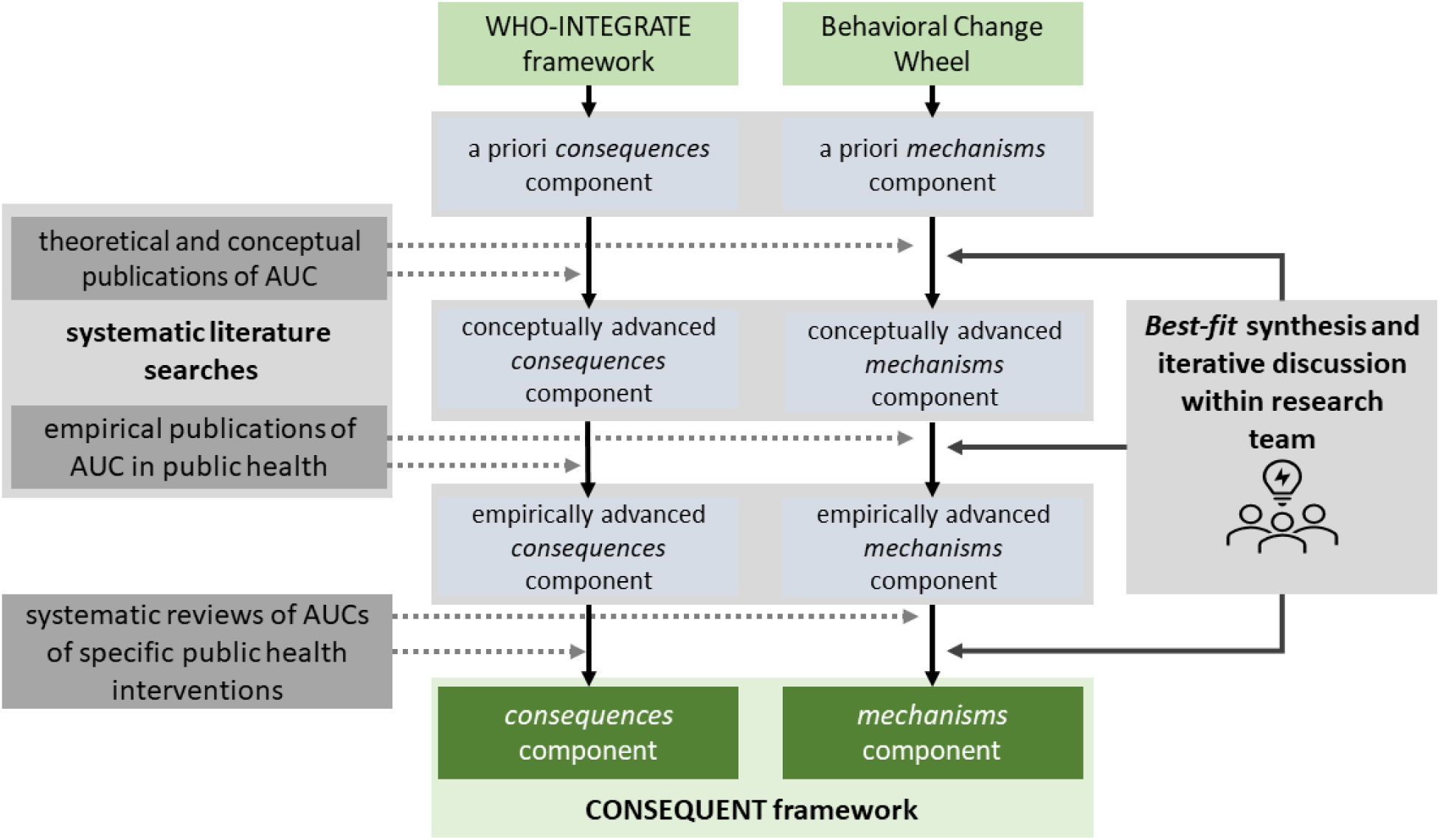
Framework development process. AUC: Adverse and other Unintended Consequences.

##### Box 1

###### Terminology

A complex (public health) intervention implemented in a complex system is likely to have diverse effects that result directly from the intervention or indirectly from its interaction with the system. In this paper, we employ the term *consequences* to define a subset of these broad system effects, and namely those, which are assigned relevance within a given normative framework. Furthermore, we employ the term *mechanisms* to refer to the direct and indirect pathways or processes through which *consequences* (beneficial or adverse) arise.

*Consequences* may be intended or unintended, depending on whether they are the outcomes the intervention is supposed to produce (29) from the perspective of those conceptualising and implementing the intervention. Further to this, consequences can be anticipated, when they are predicted in advance by intervention developers or implementers, and unanticipated, when they occur unexpectedly (29). Anticipated consequences include those that are foreseeable, i.e., where through application of analytical frameworks and experiences, consequences can reasonably be predicted in their contours, even if not in their specifics (22). Unanticipated consequences include foreseeable consequences that were not anticipated, as well as consequences (mostly in complex systems) that may not be foreseeable (e.g., black swan events) (45) and even be truly random.

Whether *consequences* are regarded as adverse (or undesirable (21) or negative (29)), beneficial (or desirable (21) or positive (29)), or neutral, depends on the perspective, the underlying normative frame and the “zoom level” (i.e. how far the effect is followed in the pathway throughout the network of effects) (29). For example, the introduction of a sugar tax leading to *a decrease in the consumption of sugar*-*sweetened beverages* may be regarded as an (intended) beneficial effect by public health practitioners, whereas it may be regarded as an adverse effect by the sugar-sweetened beverage industry. Furthermore, from a societal perspective, the intervention may lead to a dismissal of workers, increase in unemployment rates, and a loss of a source of household income. To account for the perspective-dependency in the classification of unintended consequences as beneficial or adverse, we use the overarching term adverse and other unintended consequences (AUC) in this publication.

#### 2.2.2 Development of the a priori framework

For the categorisation of *consequences*, we used the criteria and sub-criteria of the WHO-INTEGRATE framework version 1.0 (36, 46, 47). We chose this approach, as (i) it provides a reference frame that is firmly rooted in global health norms and values, as well as key public health ethics frameworks; (ii) it supports decision-making from a complexity perspective, viewing PH interventions as events in (complex) systems (6, 7, 48), and (iii) it considers outcomes of PH interventions beyond health, including social, ecological, and economic consequences.

For the categorisation of *mechanisms*, we used the Behaviour Change Wheel (40). We chose this as (i) it is the most widely used approach for examining behaviour change and (ii) it considers impacts at both individual and societal levels. We focused on the nine intervention functions in the Behaviour Change Wheel and derived a priori mechanisms based on these, including restriction, education, persuasion, incentivisation, coercion, training, enablement, modelling, and environmental restructuring.

Through brainstorming and discussions within the research team, these two frameworks were iteratively revised and advanced, resulting in the two components of the a priori framework (Supplementary Files 4 and 5).

#### 2.2.3 Identification of eligible publications for “best fit” framework synthesis

To retrieve the publications of relevance to advance the a priori framework, we conducted comprehensive literature searches in Medline (Ovid), Embase (Ovid), and the Cochrane library for systematic reviews up until November 2020. The search strategy was developed by expanding the search strategy of the 2014 scoping review by Allen-Scott and colleagues (31) and by following a guidance document by the Cochrane Adverse Effects Methods Group (49-51). In brief, the search strategy combined terms related to unintended consequences with those related to public health. The search strategy for Embase (Ovid) is provided as an example in Supplementary File 1. Additionally, we conducted forward and backward citation searches of all included studies. We conducted these searches in Scopus, Google Scholar, and Microsoft Academic.

First, to incorporate existing concepts of AUCs of PH interventions, we examined theoretical or conceptual papers which categorised, explored, or explained AUCs in-depth, grounded in or alluding to empirical findings. These included papers (i) providing *typologies or taxonomies* of AUCs of PH interventions, such as those by Allen-Scott and colleagues (31) or Lorenc and Oliver (30), (ii) describing, discussing, or exploring *mechanisms* of how PH interventions may lead to unintended consequences, such as those by Allen-Scott and colleagues (31) and Bonell and colleagues (1), as well as (iii) offering *guidance* for identifying unintended consequences of PH interventions, such as those by Bonell and colleagues (1) and Mittelmark and colleagues (52).

Second, to incorporate empirical insights to date, we retrieved and assessed systematic reviews with the primary objective to assess AUCs of PH interventions. Reviews with a primary focus on the effectiveness of interventions (i.e., the intended beneficial effects of PH interventions) were excluded.

After removal of duplicate studies, the eligibility of studies was assessed independently by two researchers (JMS, RB). Disagreements were resolved by discussion, and where necessary, by consulting with the full research team.

When considering papers for inclusion, we adopted a broad understanding of PH interventions, including a range of measures of health promotion, disease prevention, health protection, and other interventions aimed at improving health, prolonging life, and improving the quality of life among populations^64^. We excluded papers focusing on medical prevention (e.g., medical treatment of risk factors, such as hypertension), as well as secondary (e.g., prostate or breast cancer screening programmes) or tertiary prevention. Regarding borderline cases, such as vaccinations, which can be regarded as a public health or as a medical prevention effort, we excluded those publications exclusively addressing the iatrogenic effects of vaccines as medical products (e.g., fever, immune reactions), but included those reporting on the AUCs of vaccination programmes beyond adverse drug reactions (53). The full list of inclusion and exclusion criteria is provided in Additional File 2.

#### 2.2.4 Advancement of the a priori framework

As outlined above, we used the identified literature to revise the two components of the a priori framework. For this, we applied thematic analysis using a mix of inductive and deductive coding (38, 39). Specifically, the included papers were coded deductively against the categories and themes of the a priori framework, while the new themes not covered in the a priori framework were derived inductively (38, 39). The coding was done by two authors (JS, RB) using the software MAXQDA 20 (Verbi, Berlin). The thematic analysis and the framework revisions were implemented in an iterative manner (see Figure 1). The coding was conducted simultaneously for the *consequences* component and the *mechanisms* component of the framework.

First, the two components were revised and expanded based on the coding of the included theoretical and conceptual papers and the resulting new themes. The revisions were discussed in-depth within the research team, yielding conceptually advanced components. Next, the two components were further revised based on the coding of the systematic reviews of AUCs of PH interventions and discussions in the research team, yielding empirically advanced components.

#### 2.2.5 Validation of the a priori framework

To validate the comprehensiveness of the framework, four systematic reviews on unintended consequences of different PH interventions were coded against the empirically advanced framework components. We chose four different PH interventions, i.e. setting-based drug prevention (44), prevention of SARS-CoV-2 transmission in schools (41), international travel-related control measures to control COVID-19 (42), and measures to reduce the consumption of sugar-sweetened beverages (43); the systematic reviews of the AUCs of these PH interventions had been conducted by or in cooperation with research team members. After a final review and discussion within the research team, the two-component framework was finalised as the adverse and other unintended CONSEQuences of pUblic hEalth iNTerventions (CONSEQUENT) framework.

### 2.3 Results

After the removal of duplicates, the literature searches identified 2998 records. The full texts of 150 records were screened for eligibility, and 15 records met the criteria for inclusion as theoretical or conceptual publications (1, 20, 30, 31, 52, 54-61). By screening the reference lists of the included records, as well as of the identified reviews, we included another 3 records (62-64). We also identified 15 systematic reviews (9, 33, 65-76) reporting on AUCs of different PH interventions through the database searches. No additional records yielding systematic reviews were identified through searches of the reference lists. Eventually, 18 unique records of theoretical or conceptual publications and 15 unique systematic reviews were included for thematic analysis and coding. The PRISMA flow-chart visualising this process is presented in supplementary file 3 (77).

The two-component CONSEQUENT framework is presented in Figure 2.

**Figure 2:**
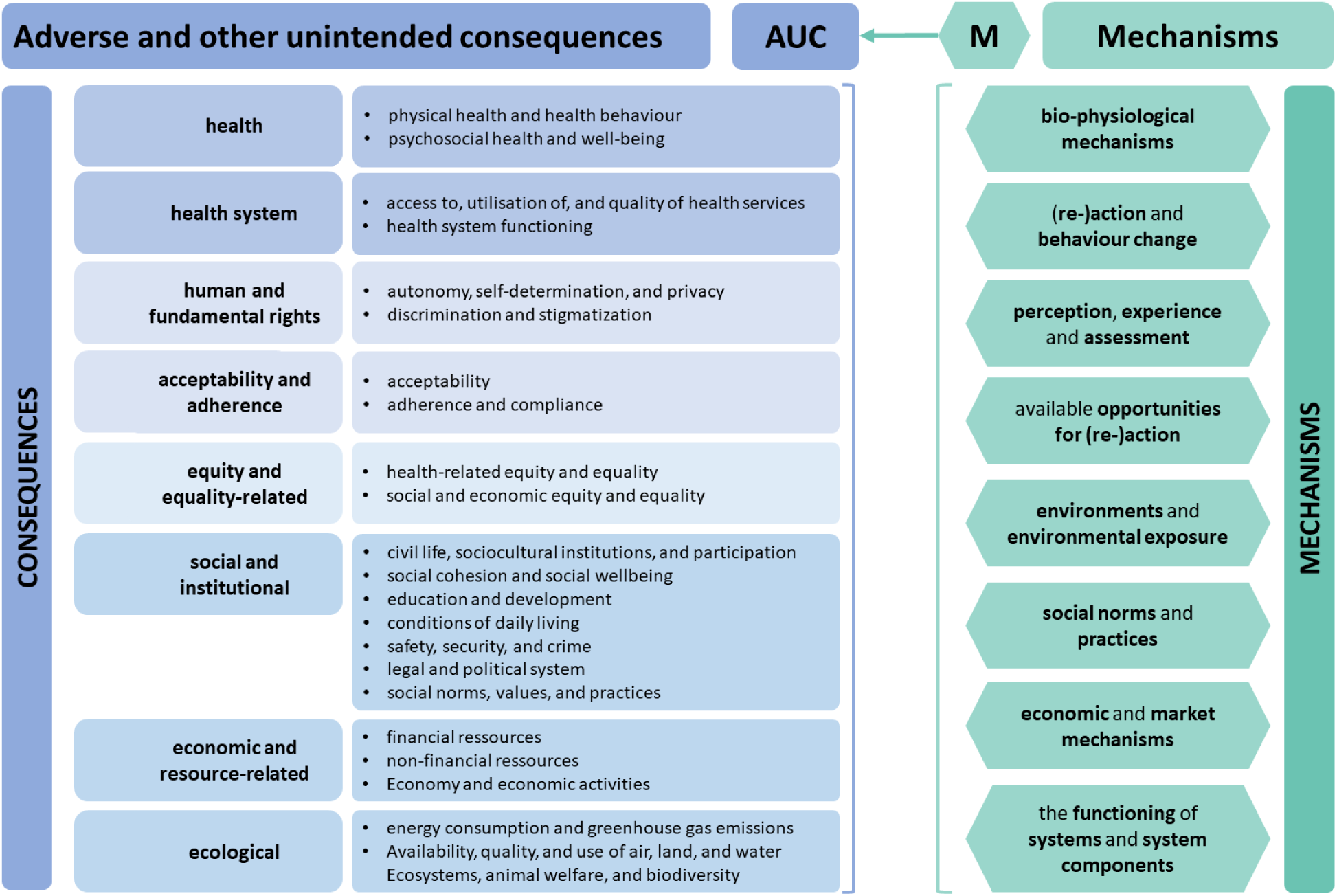
CONSEQUENT framework comprising consequences and mechanisms. AUC: adverse or other unintended consequences; M: mechanisms.

The consequences component of the CONSEQUENT framework comprises eight first-order domains: *(i) health, (ii) health system, (iii) human and fundamental rights, (iv) acceptability and adherence, (v) equality and equity, (vi) social and institutional, (vii) economic and resource-related, and (viii) ecological*. Each first-order domain also comprises several specific second-order domains. For example, the first-order domain *health* includes *consequences for physical health and health behaviour as well as psychosocial health and wellbeing* as second-order domains. Depending on the purpose and context of framework application, either the more generic first-order domains and/or the more granular second-order domains may be considered; second-order domains may also be adapted as needed (for example, differentiating the 1^st^ order domain consequence *health* in COVID-19-related and non-COVID-19 related health consequence for the assessment of a PH interventions targeting SARS-CoV-2 pandemic). Descriptions of first- and second-order domains are provided in Table 1, and further examples—in Additional File 6.

**Table 1.**
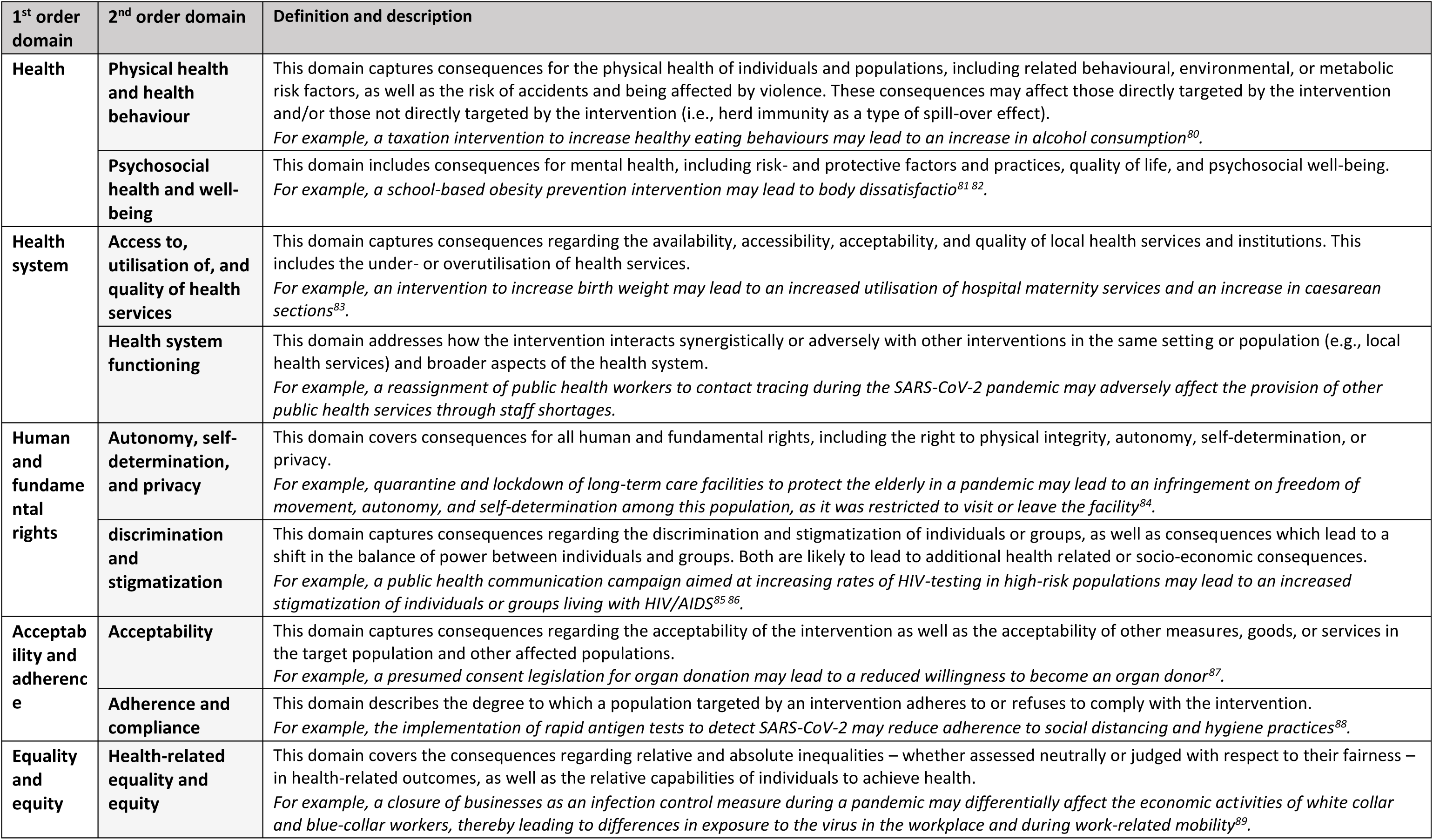
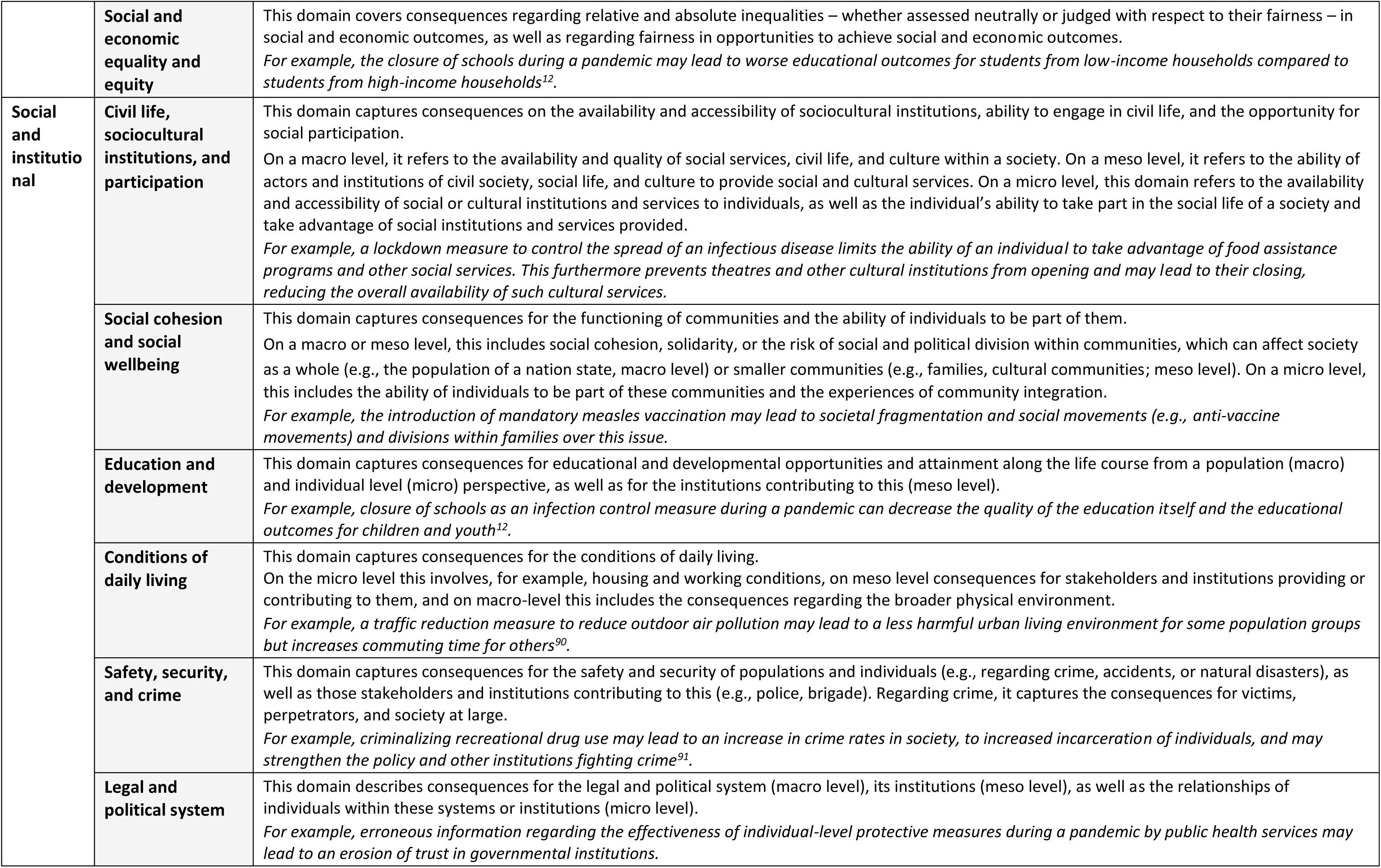
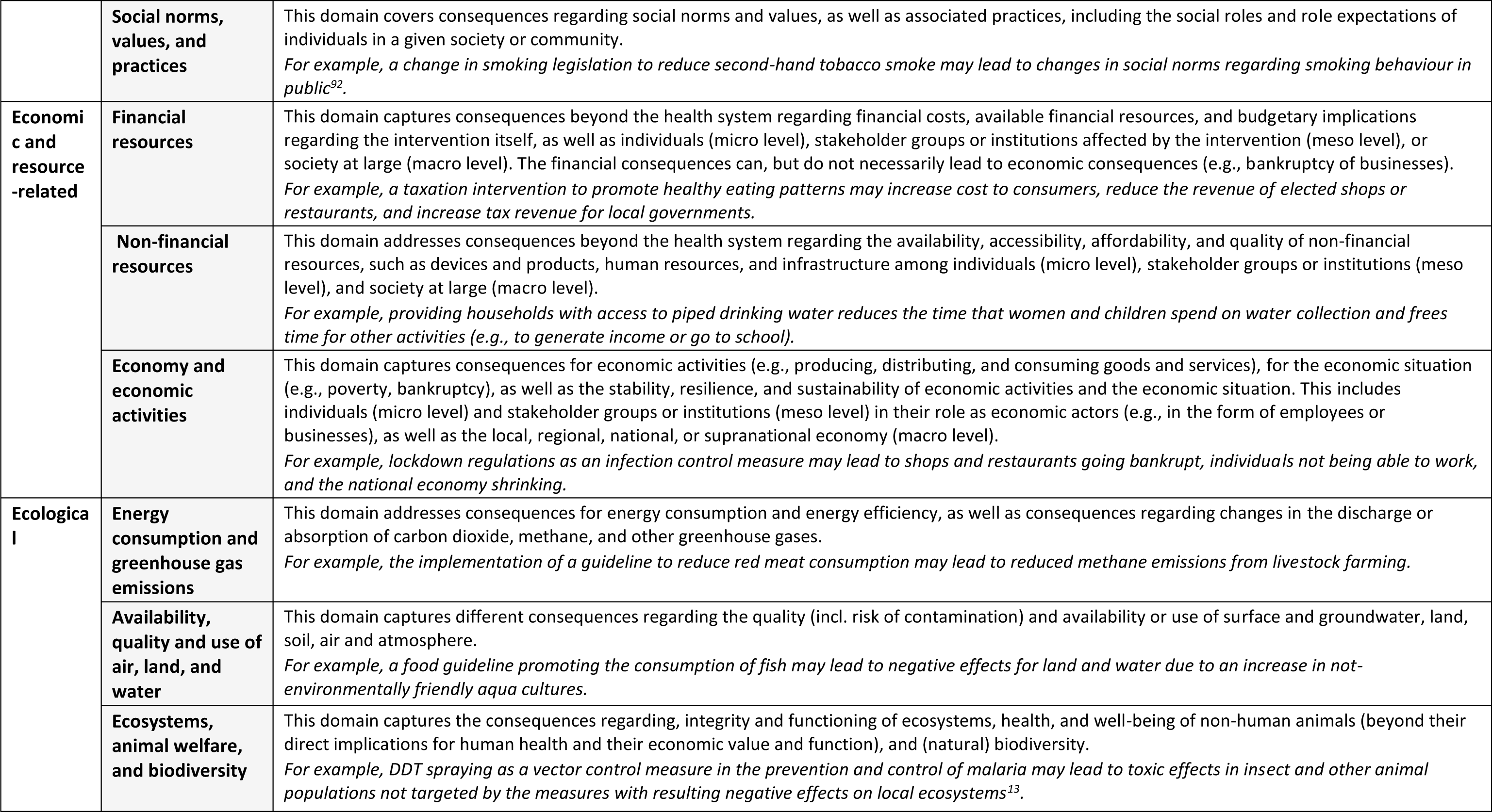
Consequences component of the CONSEQUENT framework: first-order domains, second-order domains, definition and description

| 1<sup>st</sup> order domain | 2<sup>nd</sup> order domain | Definition and description |
| --- | --- | --- |
| **Health** | **Physical health and health behaviour** | This domain captures consequences for the physical health of individuals and populations, including related behavioural, environmental, or metabolic risk factors, as well as the risk of accidents and being affected by violence. These consequences may affect those directly targeted by the intervention and/or those not directly targeted by the intervention (i.e., herd immunity as a type of spill-over effect).<br>*For example, a taxation intervention to increase healthy eating behaviours may lead to an increase in alcohol consumption<sup>80</sup>.* |
|  | **Psychosocial health and well-being** | This domain includes consequences for mental health, including risk- and protective factors and practices, quality of life, and psychosocial well-being.<br>*For example, a school-based obesity prevention intervention may lead to body dissatisfaction<sup>81 82</sup>.* |
| **Health system** | **Access to, utilisation of, and quality of health services** | This domain captures consequences regarding the availability, accessibility, acceptability, and quality of local health services and institutions. This includes the under- or overutilisation of health services.<br>*For example, an intervention to increase birth weight may lead to an increased utilisation of hospital maternity services and an increase in caesarean sections<sup>83</sup>.* |
|  | **Health system functioning** | This domain addresses how the intervention interacts synergistically or adversely with other interventions in the same setting or population (e.g., local health services) and broader aspects of the health system.<br>*For example, a reassignment of public health workers to contact tracing during the SARS-CoV-2 pandemic may adversely affect the provision of other public health services through staff shortages.* |
| **Human and fundamental rights** | **Autonomy, self-determination, and privacy** | This domain covers consequences for all human and fundamental rights, including the right to physical integrity, autonomy, self-determination, or privacy.<br>*For example, quarantine and lockdown of long-term care facilities to protect the elderly in a pandemic may lead to an infringement on freedom of movement, autonomy, and self-determination among this population, as it was restricted to visit or leave the facility<sup>84</sup>.* |
|  | **discrimination and stigmatization** | This domain captures consequences regarding the discrimination and stigmatization of individuals or groups, as well as consequences which lead to a shift in the balance of power between individuals and groups. Both are likely to lead to additional health related or socio-economic consequences.<br>*For example, a public health communication campaign aimed at increasing rates of HIV-testing in high-risk populations may lead to an increased stigmatization of individuals or groups living with HIV/AIDS<sup>85 86</sup>.* |
| **Acceptability and adherence** | **Acceptability** | This domain captures consequences regarding the acceptability of the intervention as well as the acceptability of other measures, goods, or services in the target population and other affected populations.<br>*For example, a presumed consent legislation for organ donation may lead to a reduced willingness to become an organ donor<sup>87</sup>.* |
|  | **Adherence and compliance** | This domain describes the degree to which a population targeted by an intervention adheres to or refuses to comply with the intervention.<br>*For example, the implementation of rapid antigen tests to detect SARS-CoV-2 may reduce adherence to social distancing and hygiene practices<sup>88</sup>.* |
| **Equality and equity** | **Health-related equality and equity** | This domain covers the consequences regarding relative and absolute inequalities – whether assessed neutrally or judged with respect to their fairness – in health-related outcomes, as well as the relative capabilities of individuals to achieve health.<br>*For example, a closure of businesses as an infection control measure during a pandemic may differentially affect the economic activities of white collar and blue-collar workers, thereby leading to differences in exposure to the virus in the workplace and during work-related mobility<sup>89</sup>.* |
|  | **Social and economic equality and equity** | This domain covers consequences regarding relative and absolute inequalities – whether assessed neutrally or judged with respect to their fairness – in social and economic outcomes, as well as regarding fairness in opportunities to achieve social and economic outcomes.<br>*For example, the closure of schools during a pandemic may lead to worse educational outcomes for students from low-income households compared to students from high-income households<sup>12</sup>.* |
| **Social and institutional** | **Civil life, sociocultural institutions, and participation** | This domain captures consequences on the availability and accessibility of sociocultural institutions, ability to engage in civil life, and the opportunity for social participation.<br>On a macro level, it refers to the availability and quality of social services, civil life, and culture within a society. On a meso level, it refers to the ability of actors and institutions of civil society, social life, and culture to provide social and cultural services. On a micro level, this domain refers to the availability and accessibility of social or cultural institutions and services to individuals, as well as the individual's ability to take part in the social life of a society and take advantage of social institutions and services provided.<br>*For example, a lockdown measure to control the spread of an infectious disease limits the ability of an individual to take advantage of food assistance programs and other social services. This furthermore prevents theatres and other cultural institutions from opening and may lead to their closing, reducing the overall availability of such cultural services.* |
|  | **Social cohesion and social wellbeing** | This domain captures consequences for the functioning of communities and the ability of individuals to be part of them.<br>On a macro or meso level, this includes social cohesion, solidarity, or the risk of social and political division within communities, which can affect society as a whole (e.g., the population of a nation state, macro level) or smaller communities (e.g., families, cultural communities; meso level). On a micro level, this includes the ability of individuals to be part of these communities and the experiences of community integration.<br>*For example, the introduction of mandatory measles vaccination may lead to societal fragmentation and social movements (e.g., anti-vaccine movements) and divisions within families over this issue.* |
|  | **Education and development** | This domain captures consequences for educational and developmental opportunities and attainment along the life course from a population (macro) and individual level (micro) perspective, as well as for the institutions contributing to this (meso level).<br>*For example, closure of schools as an infection control measure during a pandemic can decrease the quality of the education itself and the educational outcomes for children and youth<sup>12</sup>.* |
|  | **Conditions of daily living** | This domain captures consequences for the conditions of daily living.<br>On the micro level this involves, for example, housing and working conditions, on meso level consequences for stakeholders and institutions providing or contributing to them, and on macro-level this includes the consequences regarding the broader physical environment.<br>*For example, a traffic reduction measure to reduce outdoor air pollution may lead to a less harmful urban living environment for some population groups but increases commuting time for others<sup>90</sup>.* |
|  | **Safety, security, and crime** | This domain captures consequences for the safety and security of populations and individuals (e.g., regarding crime, accidents, or natural disasters), as well as those stakeholders and institutions contributing to this (e.g., police, brigade). Regarding crime, it captures the consequences for victims, perpetrators, and society at large.<br>*For example, criminalizing recreational drug use may lead to an increase in crime rates in society, to increased incarceration of individuals, and may strengthen the policy and other institutions fighting crime<sup>91</sup>.* |
|  | **Legal and political system** | This domain describes consequences for the legal and political system (macro level), its institutions (meso level), as well as the relationships of individuals within these systems or institutions (micro level).<br>*For example, erroneous information regarding the effectiveness of individual-level protective measures during a pandemic by public health services may lead to an erosion of trust in governmental institutions.* |
|  | **Social norms, values, and practices** | This domain covers consequences regarding social norms and values, as well as associated practices, including the social roles and role expectations of individuals in a given society or community.<br>*For example, a change in smoking legislation to reduce second-hand tobacco smoke may lead to changes in social norms regarding smoking behaviour in public<sup>92</sup>.* |
| **Economic and resource-related** | **Financial resources** | This domain captures consequences beyond the health system regarding financial costs, available financial resources, and budgetary implications regarding the intervention itself, as well as individuals (micro level), stakeholder groups or institutions affected by the intervention (meso level), or society at large (macro level). The financial consequences can, but do not necessarily lead to economic consequences (e.g., bankruptcy of businesses).<br>*For example, a taxation intervention to promote healthy eating patterns may increase cost to consumers, reduce the revenue of elected shops or restaurants, and increase tax revenue for local governments.* |
|  | **Non-financial resources** | This domain addresses consequences beyond the health system regarding the availability, accessibility, affordability, and quality of non-financial resources, such as devices and products, human resources, and infrastructure among individuals (micro level), stakeholder groups or institutions (meso level), and society at large (macro level).<br>*For example, providing households with access to piped drinking water reduces the time that women and children spend on water collection and frees time for other activities (e.g., to generate income or go to school).* |
|  | **Economy and economic activities** | This domain captures consequences for economic activities (e.g., producing, distributing, and consuming goods and services), for the economic situation (e.g., poverty, bankruptcy), as well as the stability, resilience, and sustainability of economic activities and the economic situation. This includes individuals (micro level) and stakeholder groups or institutions (meso level) in their role as economic actors (e.g., in the form of employees or businesses), as well as the local, regional, national, or supranational economy (macro level).<br>*For example, lockdown regulations as an infection control measure may lead to shops and restaurants going bankrupt, individuals not being able to work, and the national economy shrinking.* |
| **Ecological** | **Energy consumption and greenhouse gas emissions** | This domain addresses consequences for energy consumption and energy efficiency, as well as consequences regarding changes in the discharge or absorption of carbon dioxide, methane, and other greenhouse gases.<br>*For example, the implementation of a guideline to reduce red meat consumption may lead to reduced methane emissions from livestock farming.* |
|  | **Availability, quality and use of air, land, and water** | This domain captures different consequences regarding the quality (incl. risk of contamination) and availability or use of surface and groundwater, land, soil, air and atmosphere.<br>*For example, a food guideline promoting the consumption of fish may lead to negative effects for land and water due to an increase in not-environmentally friendly aqua cultures.* |
|  | **Ecosystems, animal welfare, and biodiversity** | This domain captures the consequences regarding, integrity and functioning of ecosystems, health, and well-being of non-human animals (beyond their direct implications for human health and their economic value and function), and (natural) biodiversity.<br>*For example, DDT spraying as a vector control measure in the prevention and control of malaria may lead to toxic effects in insect and other animal populations not targeted by the measures with resulting negative effects on local ecosystems<sup>13</sup>.* |

The mechanisms component of the CONSEQUENT framework, which may be treated as a supplementary component, consists of eight mechanisms (Figure 2). AUCs may arise through: *(i) bio-physiological mechanisms, (ii) (re-)action or behaviour change, (iii) perception, experience, and assessment, (iv) available opportunities for (re-)action, (v) environments and environmental exposures, (vi) social norms and practices, (vii) economic and market mechanisms, and (viii) the functioning of systems and system components*. Each mechanism also includes a non-exhaustive list of more specific processes. For example, the mechanism of (re-)action or behaviour change includes sthe following processes: (ii.a) affecting behavioural practice(s), (ii.b) evasive, resistant, or counteractive (re-)action(s) or practices, (ii.c) supplementing practices or products, (ii.d) human error or misuse, (ii.e) triggering automated behaviours, and (iii.f) lack of action or (behaviour) change. In contrast to the second-order domains of consequences, these specific processes are not intended as standalone “sub-mechanisms”, but rather illustrate how the mechanisms may operate and are likely to vary for different PH interventions. Descriptions of the mechanisms and specific processes are presented in Table 2; further details and examples are provided in Additional File 7. The relationship between the final framework and the a priori and interim versions of the framework is depicted in Additional Files 4 and 5.

**Table 2.**
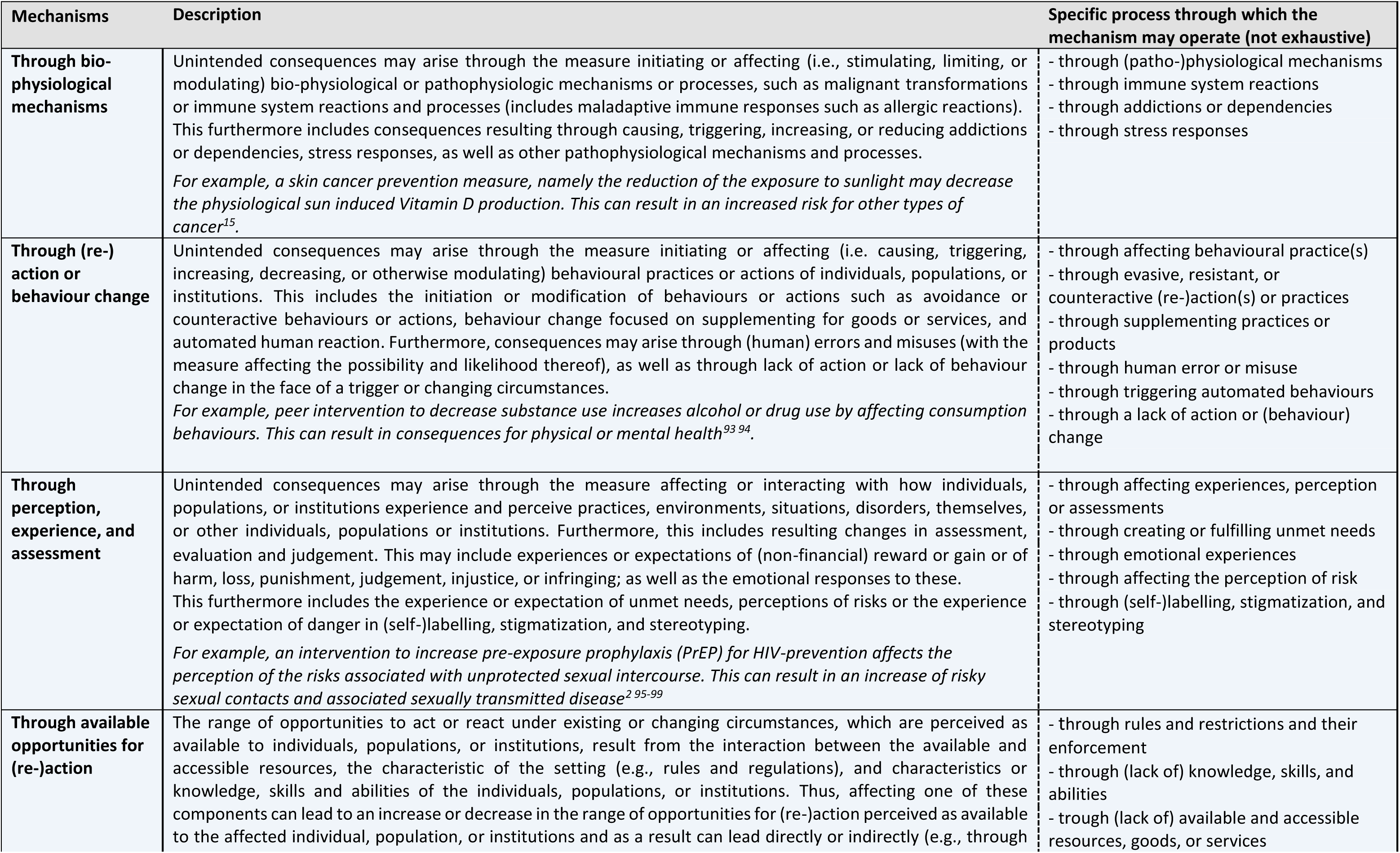
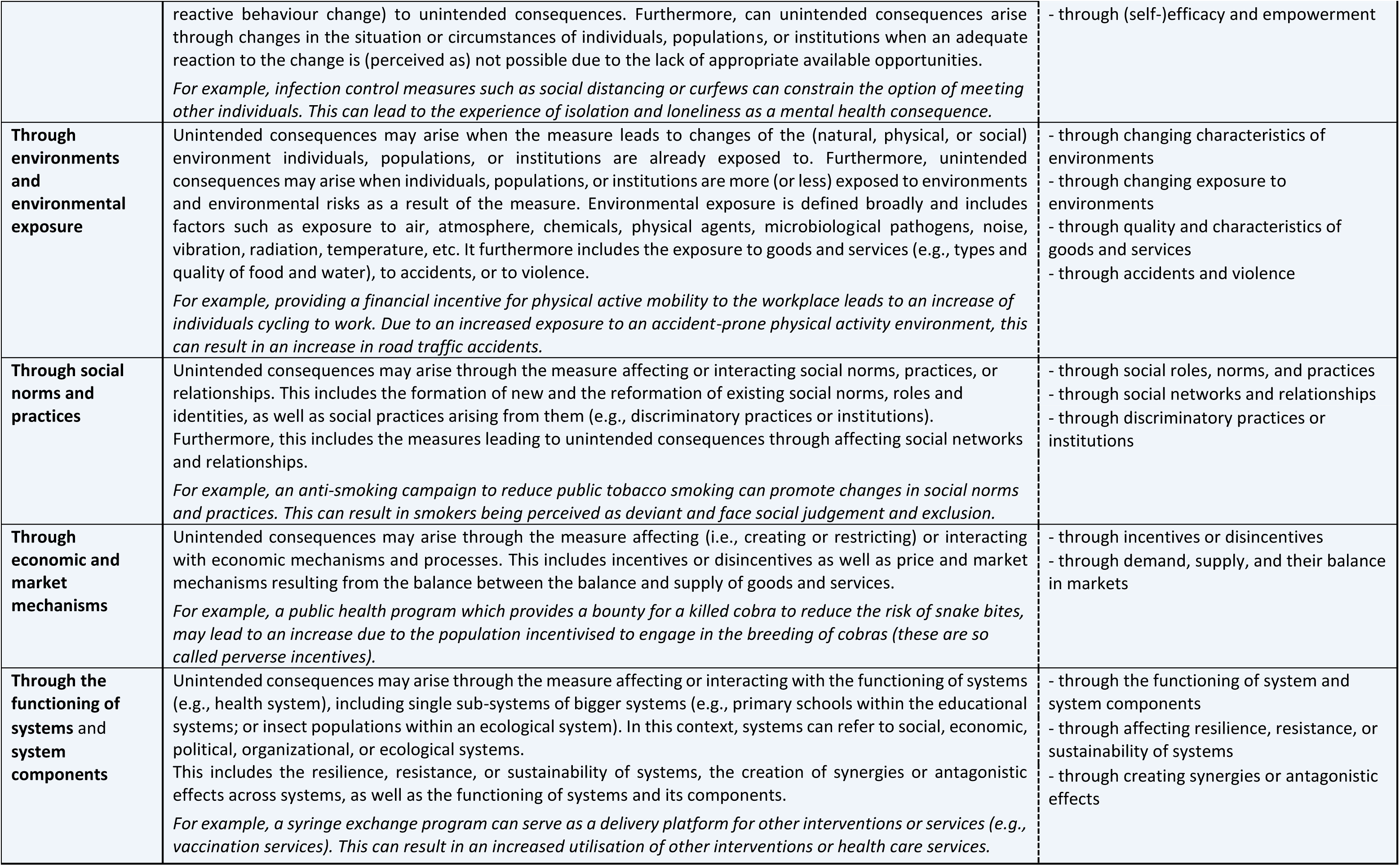
Mechanisms component of the CONSEQUENT framework: description and specific process

### 2.4 Discussion

The CONSEQUENT framework represents a novel comprehensive system to conceptualise and categorise AUCs of PH interventions, as well as the potential mechanisms leading to these. The framework is rooted in global health norms and values as it was developed drawing on the WHO-INTEGRATE framework (36); it also reflects current best insights regarding behavioural science, given its roots in the Behaviour Change Wheel (40). Furthermore, it explicitly embraces a complexity perspective (48), and thus emphasises unintended consequences of PH interventions beyond the health of individuals and populations.

#### 2.4.1 Conceptuali*s*ation of consequences and mechanisms in the CONSEQUENT framework

As shown in Figure 3, AUCs may arise through relatively simple or long and complex processes. AUCs may arise directly from the intervention (pathway A in Figure 3). For example, the taxation of sugar-sweetened beverages may lead to reduced revenue of vendors (the consequence) through a reduction in demand (the mechanism)). AUCs may also arise indirectly, when a mediator on the intended pathway leads to an unintended consequence (pathway B). For example, a public health campaign promoting physical activity may lead to an increase in road traffic injuries (consequence) due to uptake of cycling and increased exposure of cyclists to accident-prone environments (mechanism). Intended consequences may also lead to unintended consequences (pathway C). For example, skin cancer prevention programmes via a successful reduction of sun exposure (mechanism) may further lead to Vitamin D deficiencies and related health consequences (consequence) (30). Furthermore, AUCs can arise through one mechanism (pathway A) or through a combination of multiple mechanisms interacting with each other (pathway D).

**Figure 3:**
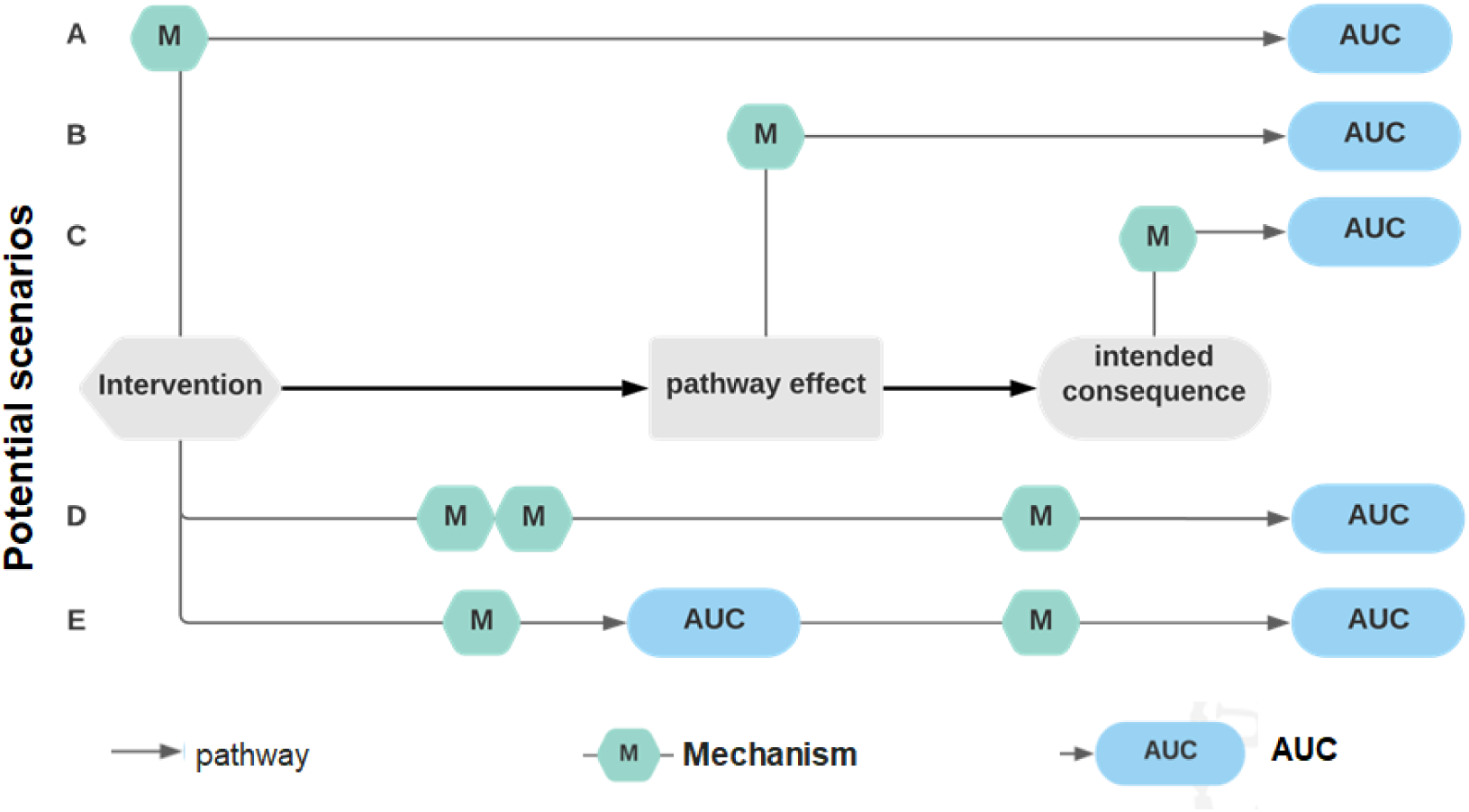
Depiction of the relations between interventions and unintended consequences. AUC: adverse or other unintended consequences; M: mechanisms.

Finally, an unintended consequence may lead to additional “secondary” unintended consequences: an unintended consequence may lead to further unintended consequences in a chain (pathway E). For example, a public health media campaign promoting healthy eating patterns may interact with and reinforce social norms and attitudes regarding obesity and obese individuals more broadly, ultimately leading to an increase in weight-based discrimination and adversely affecting the mental health of obese individuals (e.g., depression). It may also lead to lower levels of physical activity among obese individuals due to behaviours that aim to avoid further stigmatization(4, 5).

The length and complexity of the causal pathways leading to AUCs depends on the perspective of the users conceptualising these: this entails the degree to which one “zooms in” on a particular pathway. Consider the example of conceptualising the unintended consequences of a public health nutrition guideline to reduce the consumption of cholesterol. This may lead to an increase in the consumption of trans fats in margarine (change in health behaviour) and because of pathophysiological mechanisms to a further increase of cardiovascular mortality. The pathway leading to an increase in cardiovascular mortality can be adequately depicted as a long-interlinked chain of bio-physiological processes in the human body. While this conceptualisation can be helpful from a biomedical perspective, a detailed understanding of the exact chain of bio-physiological mechanisms may not be useful for public health decision-makers developing or wanting to use the public health nutrition guideline. In line with a complexity perspective (48), we suggest that the degree to which the users “zoom in” or “zoom out” on the causal pathways and thereby the level of detail considered in theorising these pathways, should be guided by the principle of usefulness for the question at hand, rather than, by the principle of comprehensiveness.

#### 2.4.2 Application of the framework

The framework was developed with two uses in mind:

The first intended use of the framework is to help public health researchers, practitioners and decision-makers conceptualise AUCs, that is, it can be used as a supporting tool to reflect on and anticipate AUCs of PH interventions when developing, evaluating, or implementing an intervention. In this application, the consequences listed in this framework are intended to guide deliberations on the potential AUCs of implementing the intervention in a given context, as well as on how different mechanisms can lead to these consequences. A comprehensive consideration of AUCs is important to appropriately judge the balance between benefits and harms of PH interventions, and anticipation of AUCs will inform their evaluation, as well as implementation of potential co-interventions or countermeasures. The CONSEQUENT framework is intended to organize these procedures and ensure that all relevant AUCs and mechanisms are considered. A simplified guidance on how to apply the framework in this conceptual manner and an illustration of this application is provided in Supplementary File 8.

The first intended use of the framework is to provide researchers with a classification system of unintended consequences of PH interventions and the mechanisms leading to them. Beyond classifying identified AUCs or mechanisms, such a classification system can reveal important gaps in the literature (41-44). For example, a preliminary version of the framework was used in a systematic review of PH interventions to prevent illicit drug use. The application of the framework showed that most publications examined in the review did not follow a structured approach for the assessment of AUCs or solely focused on health-related consequences. Furthermore, potential mechanisms were rarely described or explored. This indicated a gap in the literature on illicit drug use specifically related to the societal and ecological consequences of PH interventions for prevention (44).

#### 2.4.3 Relationship with other frameworks of intervention harms

The proposed framework shares many features with other frameworks and classification systems of the harms of PH interventions (30, 31).. We describe these below.

Allen-Scott et al. (31) propose five underlying factors of AUCs of PH interventions, such as “ignoring root causes”, “limited and/or poor quality evidence” and “lack of community engagement”. These underlying factors deviate from what we refer to as *mechanisms* in the CONSEQUENT framework. We consider the underlying factors proposed by Allen-Scott et al. to operate on a more upstream level and rather align with what we refer to as root *mechanisms*. These are understood as mechanisms operating when planning or implementing a PH intervention. Based on the publications by Allen-Scott et al (31) others (21, 23, 25), we discuss a range of relevant root mechanisms, notably: (i) not taking context into account, (ii) insufficient buy-in and participation of relevant stakeholders, (iii) (not) acting based on poor quality or insufficient information, (iv) neglecting root causes and acting based on simple answers to complex problems, and (v) (mis-) allocating scarce resources. However, more work to explore these root mechanisms is needed.

While most categories of potential harms in the framework by Lorenc and Oliver (30) are also covered in our framework, the category of “opportunity cost” is not. We did not include it in the current framework, as it requires numerous assumptions about a counterfactual reality in which the intervention would not have been implemented. However, we consider this aspect as part of the root mechanisms (i.e., “through (mis-) allocating scarce resources”).

Unintended consequences regarding equity and equality have been addressed in several publications, such as in the framework by Glover et al. for identifying and mitigating the equity harms of COVID-19 policy interventions (30, 54, 73). While these are covered in the framework component of consequences, they are not explicitly mentioned in the framework component on mechanisms as a standalone mechanism. This decision was made, as equity and inequality can arise through different mechanisms in different populations (for example, an increase in health inequality (the consequence) can arise through different populations acting within the constraints of different degrees of freedom (i.e., opportunities)).

#### 2.4.4 Strengths and limitations of the framework development process

A significant strength of the CONSEQUENT framework is the systematic, multi-pronged and iterative development of the framework. The framework has a strong and explicit normative foundation as it was modelled based on the WHO-INTEGRATE framework (36), and incorporates key insights from behavioural sciences (40). It was advanced using theoretical/conceptual, as well as empirical literature on AUCs of PH interventions derived from systematic literature searches; new insights were integrated using a mix of inductive and deductive approaches of qualitative inquiry. However, the project also has a few limitations. First, the literature searches regarding theoretical/conceptual papers and systematic reviews focusing on AUCs of PH interventions were likely not comprehensive. We conducted searches (primarily) in health-related databases; it is therefore likely that we missed insights on a broader range of consequences arising from PH interventions assessed and published by other disciplines (e.g., economics literature, environmental sciences literature). Second, the identified empirical literature itself is likely not comprehensive regarding all unintended consequences that may have occurred; for example, unintended ecological consequences were rarely addressed. Third, while we achieved content saturation in the coding process (i.e., themes were covered by multiple publications and those coded at a later stage did not provide new consequences or mechanisms), further publications may suggest additional consequences and mechanisms. For example, consideration of more publications on the AUCs of economic or market-based PH interventions derived from economic research may lead to additional insights. Finally, we focused on the literature of AUCs of PH interventions. In some cases, the distinction between economic or social policy measures and PH interventions was challenging. We aimed to overcome this issue through extensive discussions in the team and a clear definition of inclusion and exclusion criteria. For example, we are aware of the extensive literature on unintended consequences of social action from outside the field of public health. Therefore, expanding the framework based on this body of literature may provide additional insights. We therefore suggest that further validation should take place by applying it to a more diverse set of PH interventions. Based on this, a systematic collation of the experiences may lead to a further advancement of the CONSEQUENT framework, extending it into areas that are currently insufficiently covered and/or adding further granularity, such as for the second-order domains of consequences or for specific mechanisms.

## 3 Conclusion

The CONSEQUENT framework is a two-component framework to conceptualise and categorise the AUCs of PH interventions, reflecting on both outcomes (i.e., consequences), as well as the processes leading to these outcomes (i.e., mechanisms). The framework may help public health researchers, practitioners and decision-makers in conceptuali*s*ing and anticipating AUCs when developing, evaluating, or implementing public health intervention. Furthermore, the framework can be used by researchers to categori*s*e and classify AUCs of public health interventions, for example to reveal gaps in the literature on AUCs (41-44). Illustrations for the two forms of application of the CONSEQUENT Framework are provided (see Supplementary File 8) and will be further refined in future publications. Application and user-testing of the framework for practical utility may also inform further adaptations.

## Supporting information

Suppelementary Files

## Data Availability

All data produced in the present work are contained in the manuscript as well as in the public domain.

## 4 Back Matter

### 4.1 Supplementary Materials

- Supplementary File 1: Search strategy in Embase(Ovid)
- Supplementary File 2: Eligibility criteria
- Supplementary File 3: PRISMA-Flowchart
- Supplementary File 4: Relationship between the a priori and the consequences component of the final CONSEQUENT framework
- Supplementary File 5: Relationship between the a priori and the mechanism component of the final CONSEQUENT framework
- Supplementary File 6: Consequences component of the final CONSEQUENT framework with exemplary quotes
- Supplementary File 7: Mechanisms component of the final CONSEQUENT framework with exemplary quotes
- Supplementary File 8: Application of CONSEQUENT

### 4.2 Funding

The authors received funding from the German Federal Ministry of Education and Research (Bundesministerium für Bildung und Forschung; BMBF) for this project. Grant number (001001EL2032)

## 4.3 Acknowledgements

We would like to thank Peter von Philipsborn for his support in finalising the application for the funding of this project. Without his contribution, this project would likely not have been possible.

## 4.4 Author contributions

The study was conceived by JMS and EAR in close collaboration with KO and with input and support from AM and Peter von Philipsborn (LMU Munich). The search strategy was developed by JMS in close collaboration with EAR with input from AM. Literature screening and selection was conducted by JMS and RB. A preliminary coding frame was conceived by JMS drawing on discussions among all members of the research team. Application of the preliminary framework to the documents included in the analysis and the development of the revised framework versions were conducted by JMS and RB. EAR, AM, and KO reviewed and provided in-depth feedback on intermediate versions of the framework, leading to further revisions by JMS in close collaboration with RB. JMS drafted the manuscript with RB contributing specific segments. Several versions of the manuscript were critically reviewed and revised by RB, EAR, AM, and KO.

## 4.5 Institutional Review Board Statement

All study procedures were reviewed and approved by the LMU Munich institutional ethics review board. No consent to participate in the study is required.

## 4.6 Informed consent for publication

Not applicable

## 4.7 Data availability statement

The underlying data (i.e., the publications used to develop the framework) are referenced in the article and are available in the public domain. The underlying scientific publications were published in international, scientific journals. Some, although not all, are available in open access journals.

