## Supplementary material for "Anticipating and assessing adverse and other unintended consequences of public health interventions: the (CONSEQUENT) framework": Suppelementary Files

### Supplementary File 1: Search strategy in Embase(Ovid)

| | |
|---|---|
| 1 | ("harm principle" or maleficence or harmful or "side effect*" or "side-effect*").ti,ab. |
| 2 | ("harm principle" or maleficence or harmful or "side effect*" or "side-effect*").kw. |
| 3 | (harm or harmful or adverse or unintended or unintentional or Unanticipated or unwanted or paradoxical or iatrogenic) adj2 (effect or effects or reaction or reactions or event or events or outcome or outcomes or consequence or consequences)).ti,ab. |
| 4 | (harm or harmful or adverse or unintended or unintentional or Unanticipated or unwanted or paradoxical or iatrogenic) adj2 (effect or effects or reaction or reactions or event or events or outcome or outcomes or consequence or consequences)).kw. |
| 5 | ("public health" or "population health" or policy or policies or prevention or "health promotion") adj3 (program or programme or programs or programmes or intervention or interventions)).kw. or ("public health" or "population health" or policy or policies or prevention or "health promotion") adj3 (program or programme or programs or programmes or intervention or interventions)).ti,ab. |
| 6 | or/1-4<br>("harm principle" or maleficence or "side effect*" or "side-effect*").ti,ab.<br>or<br>("harm principle" or maleficence or harmful or "side effect*" or "side-effect*").kw.<br>or<br>(harm or harmful or adverse or unintended or unintentional or Unanticipated or unwanted or paradoxical or iatrogenic) adj2 (effect or effects or reaction or reactions or event or events or outcome or outcomes or consequence or consequences)).ti,ab.<br>or<br>(harm or harmful or adverse or unintended or unintentional or Unanticipated or unwanted or paradoxical or iatrogenic) adj2 (effect or effects or reaction or reactions or event or events or outcome or outcomes or consequence or consequences)).kw. |
| 8 | And/5-6 |

### Supplementary File 2: Eligibility criteria

| | **Inclusion criteria** | **Exclusion criteria** | **Comments** |
|---|---|---|---|
| Topic of publication | • Publication addresses, discusses, or explores adverse or other unintended consequences | • Publication does not address, discuss, or explore adverse or other unintended consequences | |
| Type of intervention | • Publication focuses on population-level **primary prevention or health promotion interventions** (e.g., interventions aiming to improve health of populations based on changes of health-related behaviour (e.g., handwashing) or environments (e.g., water purification) through measures such as (but not limited to) regulation, legislation, fiscal measures, environmental or social planning and (re-)structuring, or communication or marketing)<br>• **Publication focuses on (health) policy intervention** in the sense of policies, legislation or regulation aiming to improve population health (e.g., taxation of sugar-sweetened beverages, non-smoking law)<br>• We will include population level intervention and policies aiming at (health-related) harms reduction (e.g., drug legalization policies) if it explores the adverse effects of said intervention or policy (e.g., more drug deaths due to intervention intended to reduce drug deaths)<br>• We will include publications exploring a (potential) unintended consequence of PH intervention (e.g., suicide), which could at the same time be the consequence of a non-interventional process (e.g., illness such as depression), if the phenomena is clearly linked or related to a PH intervention or (health) policy<br>• We will include publications exploring the unintended effects of interventions aiming to mitigate the effects of criminal or violent behaviour | • Publication focuses on **diagnosis or treatment** of disease or health disorders and analogous **clinical interventions** (e.g., drug therapy, surgery etc.)<br>• Publication focuses on **medical prevention interventions** (e.g., medical treatment of risk factors such as hypertension)<br>• Publication focuses on **secondary prevention interventions** (e.g., clinical interventions aiming at early diagnosis and prompt treatment)<br>• Publication focuses **on tertiary prevention** (adverse effects of rehabilitation therapy after stroke).<br>• Publication focuses on **adverse drug reactions,** or the adverse effects caused by medical devices or medical procedures (e.g., surgery)<br>• Studies solely focussing on the reduction of criminal or violent behaviour (focussing on the perpetrator)<br>• Studies with interventions which do not have the primary objective of protecting nor improving health<br>• Studies which primarily focus on the medical side effects of organ donations (e.g., adverse | We aimed to include studies assessing adverse or other unintended effects of population-level public health interventions, while excluding clinical interventions, such as treatment or clinical prevention on an individual-level.<br><br>Attempt to remove studies focusing on drugs, medical devices, and medical procedures; as these are well researched and the concept that drugs, medical devices and medical procedures have adverse effects is well researched. |

| | | | |
|---|---|---|---|
| | • We will include studies which look at the unintended/adverse implications of the organ donation programs (e.g., those aiming to increase organ donation rates) | effects of the surgery or the medical criteria for eligibility)<br>• Studies investigating the adverse/unintended effects of changes in legalization / regulation of recreational drugs (though these studies will be kept for a potential further analysis/integration)<br>• Studies investigating the adverse/unintended effects of changes in the regulations for prescription opioids | |
| | | • **Screening interventions** (e.g., prostate or breast cancer screening programmes) | We will exclude population level screening interventions (e.g., new-born screening, mammography screening) as they are - in contrast to other health interventions - well researched |
| | • Primary prevention studies focusing on **vaccination** if the publication explores <u>adverse effects of vaccinations programmes</u> beyond adverse reactions against the vaccine. | • **Regarding vaccination**: publication addresses the iatrogenic effects of <u>vaccines as medical products</u> (e.g., fever, immune reactions), but does not include theoretical and/or conceptual explorations of unintended and/or adverse effects of vaccinations programmes beyond the ADR. | |
| | • Publication focuses on health system interventions | • Publication focuses on organization, management, or financing of the health care system (e.g., adverse effects of health insurance policies or task shifting)<br>• Publication focuses on interventions aiming to address availability, accessibility, affordability, acceptability or quality of diagnosis and treatment of diseases and disorders including clinical prevention interventions. | As with medical interventions, this is an area which already receives a lot of attention from areas from medicine to health economics; we therefore excluded these types of studies. |
| | AND | OR | |

| | | |
|---|---|---|
| Population / Perspective | • Publication describes an intervention aiming to improve psycho-social or physical health on a population or systems level. | • Publication describes an intervention aiming to improve mental or physical health at an individual level |
| | AND | OR |
| Type of publication | • Theoretical or conceptual publications which are discussing or exploring in-depth adverse or other unintended consequences or provide an approach to classify or systematically reflect on adverse or unintended consequences<br>• Theoretical or conceptual publication based on or rooted in empirical findings of adverse and other unintended consequences, which are explored or explained in-depth<br>• Quantitative or qualitative empirical study which explores or explains adverse or other unintended consequences in depth<br>**OR**<br>• Systematic review which explores or explains adverse or other unintended consequences in depth | • Quantitative or qualitative empirical study which merely reports on observed adverse or other unintended consequences (without deeper exploration)<br>• Systematic review of empirical studies which merely reports on observed adverse or other unintended consequences (without deeper exploration) |
| | AND | OR |
| Language | · Publication is in English, German, Spanish, Italian or French | • Publication is in languages other than English, German, Spanish, Italian, or French |
| Theoretical and conceptual studies refer to type of publication which (a) explores the *meaning* of harm, adverse or harmful consequences, or unintended consequences of public health interventions and/or provide definitions or terminologies, (b) provides *typologies or taxonomies* of adverse or other unintended consequences of public health interventions[1 2], (c) describes, explores or classifies *mechanisms* of how public health interventions led to or may lead to adverse or other unintended consequences**,** [1-3], (d) provides *domains* (e.g., domain of health, psychosocial wellbeing, individual and human right, natural environment) across which adverse or other unintended and potentially harmful consequences of public health interventions arose or may arise[4]. or (e) provides a structure, framework, or guidance to identify and/or assess adverse or other unintended consequences of public health interventions[3 5]. | | |

#### Supplementary File 3: PRISMA-Flowchart

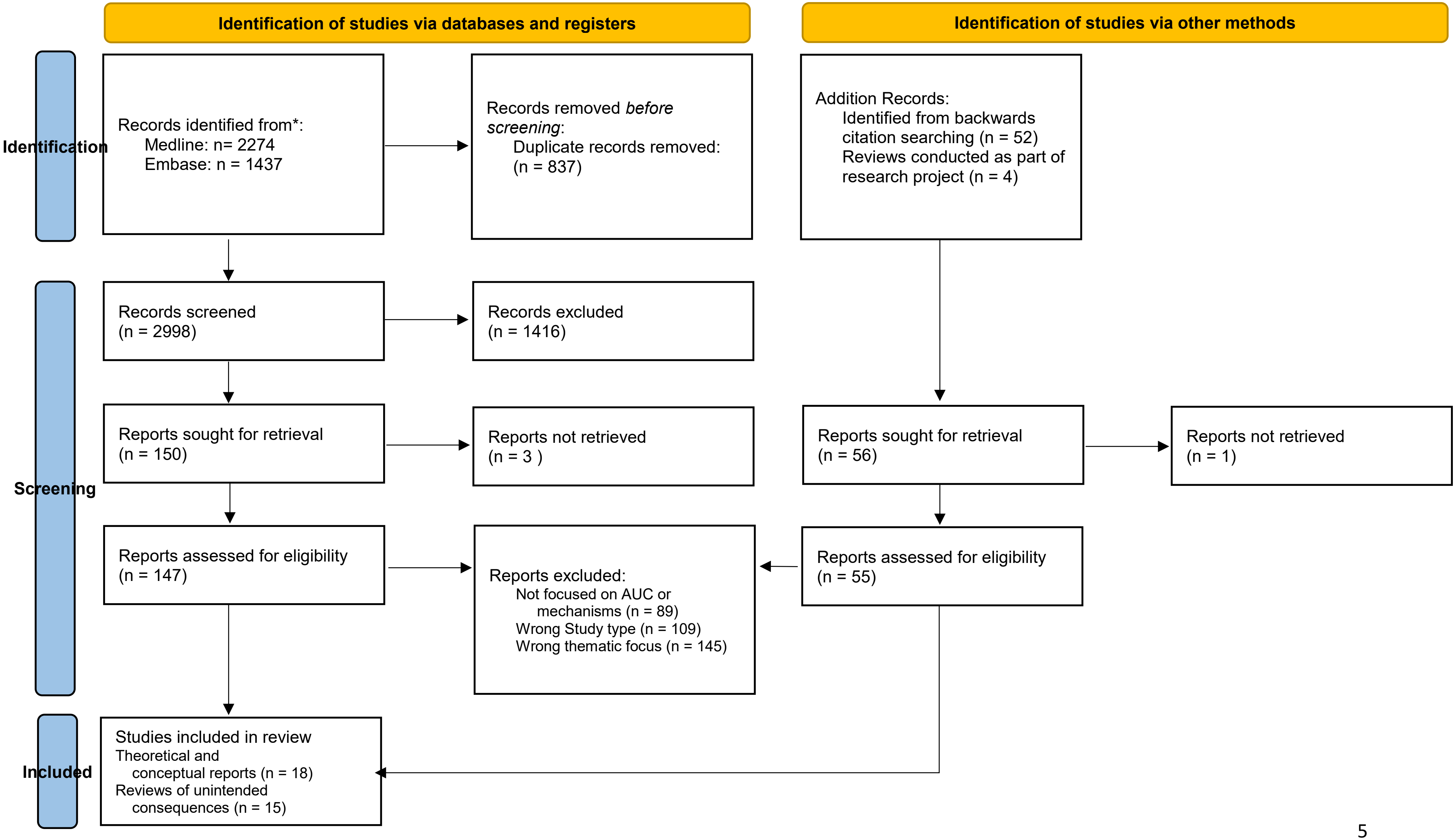

### Supplementary File 4: Relationship between the a priori and the consequences component of the final CONSEQUENT framework

| PRE-EXISTING FRAMEWORK (WHO-INTEGRATE [REF]) | | A PRIORI FRAMEWORK /THEMES | | FINAL FRAMEWORK 2$^{nd}$ ORDER DOMAINS | | FINAL FRAMEWORK 1$^{st}$ ORDER DOMAINS |
|---|---|---|---|---|---|---|
| Health-related consequences | ► | Physical health and well-being | ► | **Physical health and well-being** | ► | **Health-related consequences** |
| | | Psychosocial health and well-being | | **Psychosocial health and well-being** | | |
| Human and fundamental rights consequences | ► | Human and fundamental rights | ► | **Human and fundamental rights** | ► | **Human and fundamental rights consequences** |
| | | | | **Discrimination and stigmatization** | | |
| Consequences related to acceptability and adherence | ► | Acceptability | ► | **Acceptability** | ► | **Consequences related to acceptability and adherence** |
| | | Adherence and compliance | | **Adherence and compliance** | | |
| Consequences for equality and equity | ► | Health-related equality and equity | ► | **Health-related equality and equity** | ► | **Consequences for equality and equity** |
| | | Socio-economic equality and equity | | **Socio-economic equality and equity** | | |
| Societal implications | ► | Social outcomes and participation | ► | **Civil life, sociocultural institutions, and participation** | ► | **Social and institutional consequences** |
| | | | | **Social cohesion and social wellbeing** | | |
| | | | | **Consequences for education and personal development** | | |

| | | | | | | |
|---|---|---|---|---|---|---|
| | | Communities and social cohesion | | **Consequences for the conditions of daily living** | | |
| | | | | **Consequences for safety and security** | | |
| | | Social norms and values | | **Consequences for the legal and political system** | | |
| | | | | **Social norms, values, and practices** | | |
| | | Financial consequences | | **Financial consequences** | | |
| Financial and economic considerations | ► | Resource-related consequences | ► | **Resource-related consequences** | ► | **Economic and resource-related consequences** |
| | | Economic consequences | | **Economic consequences** | | |
| Health system consequences | ► | Access to and utilisation of healthcare | ► | **Access to and utilisation of healthcare** | ► | **Health system consequences** |
| | | Health system functioning | | **Health system functioning** | | |
| | | Energy consumption and greenhouse gas emissions | | **Energy consumption and greenhouse gas emissions** | | |
| Environmental consequences | ► | Availability and quality of air, land, and water | ► | **Availability and quality of air, land, and water** | ► | **Environmental consequences** |
| | | Animals, ecosystems, and biodiversity | | **Animals, ecosystems, and biodiversity** | | |

### Supplementary File 5: Relationship between the a priori and the mechanism component of the final CONSEQUENT framework

| PRE-EXISTING FRAMEWORK (BCW[1]) | A PRIORI FRAMEWORK / THEMES | FINAL FRAMEWORK | SPECIFIC DIMENSIONS INCLUDED IN THE MECHANISMS |
|---|---|---|---|
| BHW-framework in general | Through changing behaviours | **Through action or behaviour change** | ***Through lack of action or behaviour change*** |
| | | | ***Through evasion, resistance, or counteracting*** |
| | | | ***Through replacing or supplementing practices or products*** |
| | | | ***Through human error in or misuse*** |
| | | | ***Through automated behaviours*** |
| | Through affecting metabolic or physiological reactions | **Through bio-physiological mechanisms** | ***Through addictions or dependencies*** |
| | | | ***Through stress responses*** |
| Restrictions | Through restricting practices, goods, or services | **Through available opportunities** | ***Through rules and restrictions and their enforcement*** |
| Enablement | Through empowering individuals and communities | | ***Through (self-)efficacy and empowerment*** |
| Education and training | Through increasing knowledge or understanding | | ***Through available and accessible resources, goods, and services*** |
| Training | Through improving skills and abilities | | ***Through knowledge, skills, and abilities*** |
| Persuasion | Through changing perceptions and attitudes | **Through experiences or perceptions** | ***Through unmet needs*** |
| | | | ***Through emotional experiences*** |
| | Through affecting psychological reactions | | ***Through the perception of risk*** |
| | | | ***Through (self-)labelling, stigmatization, and stereotyping*** |

| | | | |
|---|---|---|---|
| Incentivisation and coercion | Through providing financial, economic, or social incentives (or disincentives) | **Through economic mechanism** | ***Through incentives or disincentives***<br>***Through demand, supply, and their balance in markets*** |
| Environmental restructuring (environmental context) | Through changing environmental exposures<br>Through triggering automated responses<br>Through changing the availability, accessibility, or quality of goods or services | **Through environments and environmental exposure** | ***Through quality and characteristics of goods and services***<br>***Through quality and characteristics of goods and services*** |
| Environmental restructuring (social context)<br><br>Modelling | Through changing social practices and norms | **Through social norms and practices** | ***Through affecting social role, norms, and practices***<br>***Through social networks and relationships***<br>***Through discriminatory practices or institutions*** |
| | **New identified themes** | **Through the functioning of systems** | ***Through resilience, resistance, or sustainability of systems***<br>***Through creating synergies or antagonistic effects***<br>***Through affecting the functioning of system components*** |

### Supplementary File 6: Consequences component of the final CONSEQUENT framework with exemplary quotes

| 1st order domain | 2nd order domain | Exemplary passages |
|---|---|---|
| **Health** | **Physical health and health behaviour** | „Physical harm, meaning a harm occurring to the physical structure of a human who was associated with a PH intervention [public health interventions], was the most commonly occurring PH intervention associated harm in this study (n = 20). Infants and children had a disproportionally greater risk of experiencing physical harms associated with PH intervention in the areas of: birth weight, obesity, food supplementation, infectious diseases, and suicide as these PH intervention are most often targeted at this population (Table 1).“ (Allen-Scott 2014[2]) |
| | **Psychosocial health and well-being** | “Burgess, Fu and van Ryn review literature showing that some mothers who are exposed to health education about the harm to themselves and their children because of their smoking, unfortunately experience lower self worth and increased guilt and sadness.” (Mittelmark 2014[3]) |
| **Health system** | **Quality of, access to and utilisation of health services** | “In the context of a public health crisis, stigmatized individuals may avoid treatment and testing because they fear being exposed to the judgmental attitudes of medical professionals who they believe are likely to hold negative feelings concerning them“. (Quinn 2018[4]) |
| | **Health system functioning** | “It [the harm reduction approach] justifies proffering information and services to help individuals avoid certain risks even if this appears to condone practices judged by society as anti-social or even immoral. Proponents of this approach often refer not only to the obligation to help people with special needs to avoid serious harm, but also to the utility of this approach. For example, syringe-exchange programmes for injection drug users can serve as delivery sites for other health services, and help gain the trust of this under-served population.**“** (Guttman 2004[5]) |
| **Human and fundamental rights** | **Autonomy, self-determination, and privacy** | “Those seeking HIV prevention and treatment face the risk of stigma, discrimination and human rights abuses.” (Allen-Scott_2014)<br><br>“Diskutiert werden [bezüglich der Auswirkungen von Gesundheitstechnologien] unter anderem unerwünschte Effekte durch: […] Missachtung von Persönlichkeitsrechten, Datenzusammenführungen und bewusste Angriffe durch Dritte.“ (Schütz 2020[6]) |
| | **Discrimination and stigmatization** | “Whether the information that is provided is accurate or not it may lead to problems such as stigmatisation and discrimination for vulnerable groups. [...] For individuals who are stigmatised, the consequences can be dramatic. In the context of a public health crisis, stigmatized individuals may avoid treatment and testing because they fear being exposed to the judgmental attitudes of medical professionals who they believe are likely to hold negative feelings concerning them.“ (Quinn 2018[4]) |

| | | |
|---|---|---|
| **Acceptability and adherence** | **Acceptability** | "The emission of incorrect information will furthermore result in an erosion of trust of the public authorities concerned, reducing their ability to be effective in future crisis" (Quinn 2018[4]) |
| | **Adherence and compliance** | "Among substance abuse patients treated in the Department of Veterans Affairs, compared with patients who remained stable or improved, patients who deteriorated saw their treatment program as less supportive and expressive and less oriented toward self-understanding. These findings illustrate the potential link between poor treatment alliance and deterioration in treatment" (Guttman 2004[5]) |
| **Equality and equity** | **Health-related equality and equity** | "Health communication interventions, particularly those that are successful, may reinforce, rather than reduce, existing social disparities.60 Research findings indicate that, following the dissemination of health information, populations from higher socio- economic groups were more likely to have increased knowledge relevant to the health issue and more likely to adopt recommended practices, though motivation to do so may have been similar across different populations. This phenomenon is called the 'knowledge gap' and it may not be ethically problematic in commercial contexts, but is an ethical problem in public health." (Guttman 2004[5]) |
| | **Social and economic equality and equity** | "interventions may create harm by worsening health inequalities. That is, some successful interventions may improve outcomes across the population, but exacerbate existing inequalities by benefiting privileged groups more than disadvantaged groups. [...] It has also been argued that targeted or 'high-risk' approaches are more likely than population-level initiatives to widen health inequalities." (Lorenc 2014[7]) |
| **Social and insitutional** | **Civil life, sociocultural institutions, and participation** | "In addition to the harms that occur to individuals, groups and their interests in society, incorrect communication also represents a waste of public resources that could be used more effectively. The emission of incorrect information will furthermore result in an erosion of trust of the public authorities concerned, reducing their ability to be effective in future crisis." (Quinn 2018[4])<br>„A cultural harm refers to any damage to a population's ''way of life'', which includes language, arts and sciences, spirituality, social activity and interactions (RCHI 2013)." (Allen-Scott 2014[2]) |
| | **Social cohesion and social wellbeing** | "More indirect psychological harms may result where targeted health behaviours are bound up with individuals' social identity or relationships with others. […] The qualitative literature suggests that, for example, moderate consumption of alcohol or other drugs may facilitate social interactions,15 16 and that unprotected sex may facilitate trust and intimacy within sexual relationships.17 18 It is thus possible that interventions targeting these behaviours could have negative impacts on social or intimate relationships." (Lorenc 2014[7]) |
| | **Education and development** | "Two articles described evaluations of the Reconnecting Youth program that was designed to decrease school deviance and involvement with drugs and improve management of moods for youth exhibiting high-risk behaviors. A study by Cho, Hallfors, and Sanchez (2005) found that at 6-month follow-up, only negative program effects were seen; students showed decreased GPA, decreased school connectedness, lower conventional peer-bonding, increased anger, and higher affiliation with high-risk peers." (Kuiper 2018) |

| | | |
|---|---|---|
| | **Conditions of daily living** | No suitable paragraph was found. Derived theoretically by discussion within the research team. |
| | **Safety, security, and crime** | "A potential negative effect of tobacco tax is its impact on illegal cross-border transport and sale of untaxed tobacco products. […] The World Bank considers that ''rather than foregoing [tobacco] tax increases, the appropriate response to smuggling is to crack down on criminal activity''." (Wilson 2005[8]) |
| | **Legal and political system** | "The emission of incorrect information will furthermore result in an erosion of trust of the public authorities concerned, reducing their ability to be effective in future crisis." (Quinn-2018[4]) |
| | **Social norms, values, and practices** | "In contrast, evaluations of some drug-prevention interventions have noted adverse effects on the use of drugs and provided evidence that these may be mediated by interventions bringing recipients into contact with more risk-involved peers and reinforcing pro-risk attitudes and behaviours." (Bonnell 2015[9]) |
| **Economic and resource-related** | **Financial resources** | "The households of smokers who succeed in quitting after a tax increase will save on tobacco-related spending, through lower out-of-pocket medical expenses, lower health and life insurance premiums, and lower cleaning costs for clothes and furnishings. For a society, there is a direct financial benefit from tobacco tax when some of the tax revenue is tied to tobacco control spending, as it can assist in funding programmes that reduce smoking prevalence (eg., CDC (Centers for Disease Control and Prevention), 1996). Less directly, any reduction of smoking attributable to taxation levels should reduce health sector costs overall." (Wilson 2005[8]) |
| | **Non-financial resources** | "A reduction in tobacco sales from tobacco taxation might be expected to reduce employment in tobacco production, cigarette manufacture, and tobacco sales. In the long term however, new jobs should be created elsewhere in national economies as a result of re-diverted consumer spending (Jha & Chaloupka, 2000; Warner, 2000). The majority of workers would probably benefit in the long-term, since economies should grow more as tobacco sales decline (since tobacco use impairs work- force productivity and reduces the size of the population of consumers)." (Wilson 2005[8]) |
| | **Economy and economic activities** | "An economic harm, defined as damage that relates to production, distribution and consumption of goods and services, was identified in four of the included studies. [...] Balog (2009) reported that if the long-term effects of a vaccine are unknown, individuals, governments and private companies might waste resources to rollout this vaccine with no long-term benefits for the population." (Allen-Scott 2014[2]) |
| **Ecological** | **Energy consumption and greenhouse gas emissions** | "It is estimated that transport-related fuel use accounts for around 22% of CO2 fuel emissions. Although individual fuel use may have a negligible impact, an accumulation of increased fuel emissions may have significant environmental, economic and health impacts at a global level." (Thomson 2008[10]) |

| | | |
|---|---|---|
| | **Availability, quality and use of air, land and water** | No suitable paragraph was found. Derived theoretically by discussion within the research team. |
| | **Ecosystems, animal welfare and biodiversity** | No suitable paragraph was found. Derived theoretically by discussion within the research team. |

### Supplementary File 7: Mechanisms component of the final CONSEQUENT framework with exemplary quotes

| Mechanism | Dimension | Description | Example |
|---|---|---|---|
| **Through bio-physiological mechanisms** | Definition and description | Unintended consequences may arise through the measure initiating or affecting (i.e., stimulating, limiting, or modulating) bio-physiological or pathophysiologic mechanisms or processes, such as malignant transformations or immune system reactions and processes (includes maladaptive immune responses such as allergic reactions).<br>This furthermore includes consequences resulting through causing, triggering, increasing, or reducing addictions or dependencies, stress responses, as well as other pathophysiological mechanisms and processes.<br>*For example, a skin cancer prevention measure, namely the reduction of the exposure to sunlight may decrease the physiological sun induced Vitamin D production. This can result in an increased risk for other types of cancer[11].* | *"If parasites affect school performance (for example, by reducing emetic iron, causing weakness and inattention), the treatment may result in both treated and untreated children learning more." (Angelucci 2015[12])* |
| | Through addictions or dependencies | Unintended consequences may arise through the measure initiating or affecting addictions or dependencies in individuals or populations. This includes chemical as well as behavioural addictions, as neurophysiologic reinforcement is seen as a primary factor in the development of addictions and dependencies. Furthermore, withdrawal reactions to the lack of the addictive substance following prohibition as a public health measure are included as well.<br>*For example, a public health measure to prohibit alcohol consumption in homeless shelters may lead to hospitalizations due to withdrawal*. | *No suitable paragraph was found. Derived theoretically by discussion within the research team.* |
| | Through immune system reactions | Consequences may arise through the measure affecting or interacting immunologic reactions to exposures or the lack thereof. This includes maladaptive immune responses such as allergic reactions as well as the onset of inflammatory diseases.<br>*For example, if the lack of exposure to peanuts in high-risk children as a public health measure leads to higher rates of peanut allergies in the intervention group[13].* | *"There are complex interactions between human genetic factors and this changing environment that is leading to the increasing prevalence of metabolic and inflammatory diseases. Alterations to human gut bacterial communities (the microbiota) and lowered prevalence of helminth infections are potential environmental factors contributing to immune dysregulation." (Stiemsma 2015[14]).* |

| Mechanism | Dimension | Description | Example |
|---|---|---|---|
| | Through pathophysiology | Consequences may arise through the measure initiating and / or affecting pathophysiologic mechanisms or processes. This included malignant transformations as well as other pathological processes like atherosclerosis. *For example, a public health message to reduce cholesterol in the diet leads to an increase in the consumption of margarine, which increases the risk of cardio-vascular mortality due to the bio-pathological reaction of the human body to the exposure to trans-fats.* | |
| | Through stress responses | Unintended consequences may arise through the measure affecting acute and chronic stress. This includes the bio-physiological reaction to stressors. *For example, a public health measure to promote healthy eating and physical activity might cause obese individuals targeted by these measures to experience stress, which can effect multiple physical and mental health outcomes (e.g., anxiety, eating disorders)[15-17].* | *"With regard to the first mechanism, the epidemiologist Michael Marmot has argued that many of the social determinants of health like education, income, and social capital create health disparities through a common pathway; namely, chronic stress. Individuals who are less educated, poor, and with limited social capital have less control over their lives, which leads to stress and contributes to poor cardiovascular health." (Courtwright 2009[18])* |
| **Through action or behaviour change** | Definition and description | Unintended consequences may arise through the measure initiating or affecting (i.e. causing, triggering, increasing, decreasing, or otherwise modulating) behavioural practices or actions of individuals, populations, or institutions. This includes the initiation or modification of behaviours or actions such as avoidance or counteractive behaviours or actions, behaviour change focused on supplementing for goods or services, and automated human reaction. Furthermore, consequences may arise through (human) errors and misuses (with the measure affecting the possibility and likelihood thereof), as well as through lack of action or lack of behaviour change in the face of a trigger or changing circumstances. *For example, peer intervention to decrease substance use increases alcohol or drug use by affecting consumption behaviours. This can result in consequences for physical or mental health[19 20].* | *"Consequently, VNPs [vaporized nicotine products] could reduce harm to never smokers who would have otherwise initiated long-term cigarette use, and reduce harm to current smokers by helping them to quit, to switch to exclusive VNP use or to substantially reduce their smoking." (Levy 2017[21])* |
| | Through lack of action or behaviour change | Unintended consequences may arise through the measure not initiating or affecting actions or behavioural change. This includes unintended consequences resulting from the stringency and timing of action or behaviour change (e.g., delay of seeking medical help). *For example, if in the context of a rapidly developing pandemic, government and public health decision makers delay the implementation of non-pharmacological anti-pandemic measures, this can lead to an additional increase in the number of infections or deaths[22].* | *"In healthcare settings, women who perceive stigmatization from their providers report delaying use of preventive health services for fear of being judged or embarrassed. This avoidance of care allows for untreated problems to progress to a more advanced stage that may be more difficult to treat, thus exacerbating health problems." (Stangl 2019[23])* |

| Mechanism | Dimension | Description | Example |
|---|---|---|---|
| | Through evasion, resistance, or counteracting | Consequences may arise through the measure affecting or interacting with the levels of adherence to the measure or rules and regulations, as well as attempts to evade and circumvent or resist and counteract them.<br>*For example, if attempts to undermine or circumvent an alcohol tax through engaging in home-made beer brewing could lead to adverse effects due to exposure to harmful products without quality control (e.g., iron intoxication through beer; methanol intoxications)[24].* | *"Action: Tax sugar-sweetened beverages (SSBs). Documented unintended consequence: Increased consumption of beer ,beyond the decrease in sugar-sweetened beverages. (Brown 2013[25])* |
| | Through supplementing practices or products | Unintended consequences may arise through the measure leading to or interacting with behaviour change or action in the form of practices or products being replaced by other products or practices which serve as supplements, or which are intended to compensate for a lost function. This includes substituting or compensating behaviour as well as effects triggered through characteristics of the substitutes.<br>*For example, if sugar taxes reduce the consumption of sugar sweetened beverages (SSBs), but the consumption is replaced by alcoholic beverages such as beer, serving as a supplement, leading to an increase in alcohol consumption[26].* | *"It cannot be assumed that a shift from car use to more physically active forms of transport will automatically lead to an increase in overall levels of physical fitness or activity. For example, gym exercise may be replaced by cycling to work. However, one study assessed changes in fitness among those who changed from driving to walking or cycling to work; levels of fitness and walking speed improved" (Thompson 2008[10]).* |
| | Through human error in or misuse | Unintended consequences may arise though the measure affecting the probability of human error or misuse of practices, goods, or services. This also includes the measures affecting the severity of the consequences of human error or misuse, or affecting the presence and effectiveness of institutions preventing human error or misuse and mitigating their consequences.<br>*For example, if public health information recommends practicing the calendar-method as a method for birth control. While the method is effective in protecting against unwanted pregnancies when applied correctly, unintended consequences may arise from human error, which is likely to occur[27].* | *"Before we can discuss the effectiveness of any contraceptive method, the distinction between method failure and user failure must be clear. When a contraceptive method fails because of a defect in the product itself, it is method failure. For example, if a condom breaks because of an inherent weakness in the latex, it is a method failure. When it fails because of incorrect or inconsistent use, it is user failure. For example, if a condom breaks, leaks, or slips off because the user puts it on incorrectly, it is a user failure. The combination of both is called contraceptive failure " (Haignare 1999[27])* |
| | Through triggering automated behaviours | Unintended consequences may arise through the measure initiating or affecting automated behaviours, including related aspects of self-control or self-regulation.<br>*For example, after exposure to a public health measure which leads an overweight and obese individual experience stigma, they may feel less able to control their eating behaviour and eat a much larger amount of high-calorie snack food than intended[28].* | *"Numerous studies have shown that, for members of stigmatized groups, situations that trigger expectations or concerns that one will be stigmatized impair performance and diminish self-regulatory processes. This suggests that stigmatization may lower regulation processes needed to make changes in one's health behaviour, including, but not limited to, smoking." (Burgess 2009[29])* |

| Mechanism | Dimension | Description | Example |
|---|---|---|---|
| **Through experiences and judgements** | Definition and description | Unintended consequences may arise through the measure affecting or interacting with how individuals, populations, or institutions experience and perceive practices, environments, situations, disorders, themselves, or other individuals, populations or institutions. Furthermore, this includes resulting changes in assessment, evaluation and judgement. This may include experiences or expectations of (non-financial) reward or gain or of harm, loss, punishment, judgement, injustice, or infringing; as well as the emotional responses to these.<br>This furthermore includes the experience or expectation of unmet needs, perceptions of risks or the experience or expectation of danger in (self-)labelling, stigmatization, and stereotyping.<br>*For example, an intervention to increase pre-exposure prophylaxis (PrEP) for HIV-prevention affects the perception of the risks associated with unprotected sexual intercourse. This can result in an increase of risky sexual contacts and associated sexually transmitted disease[30-35].* | *"When consumers view a food warning, they may experience a range of short-term negative emotional responses [..] for example, consumers may feel fear and anxiety in response to the knowledge that a product contributes to health harms. [...] Evidence suggests that experiencing anxiety, fear, or other negative emotions in response to warnings is a productive reaction because these emotions are a key pathway through which warnings encourage healthier behaviours and promote informed choice." (Grummon 2020[36])* |
| | Through unmet needs | Unintended consequences may arise through the measure affecting or interacting with the experience or expectation of unmet needs, which may be created by unavailability of a practice, institution, or good, which serves a particular function, no longer being available.<br>*For example, smokers quitting or lowering tobacco consumption because of an increase in taxation, may lead to the unmet need of measures for stress relief, through the practice of smoking breaks no longer being available[8].* | *"Certain practices, such as smoking, might offer members of vulnerable groups not only pleasure, but also important coping mechanisms or serve social functions that are not easily replaced. The less privileged economically are also likely to have fewer options for healthier substitutions for practices they enjoy that are considered unhealthy." (Guttman 2004[5])* |
| | Through emotional experiences | Unintended consequences may arise through the measure affecting a range of emotions such as anger, guilt, disgust, shame, uncertainty, dissonance, or distress, or the anticipation thereof. Those can result from experiences or expectations of reward or things such as loss, punishment, judgement, or injustice.<br>*For example, a public health campaign to promote healthy eating and physical activity may lead to body shaming and or distress in individuals not conforming to a certain body norm[15 16 23].* | *"The dissemination of health messages through educational or media campaigns may generate damaging feelings of worry or guilt, which can have negative effects not only on general well-being but, in many cases, on the targeted behaviours themselves. While most media campaigns do now try to avoid explicitly guilt-oriented messages, there is still considerable potential for harms, which have rarely been investigated systematically." (Lorenc 2014[11])* |
| | Through the perception of risk or experience and expectation of danger | Unintended consequences may arise through the measure affecting or interacting with perceptions of risks or the experience or expectation of danger. This includes the perception and experience of the probability of occurrence as well as the severity of consequences. This furthermore includes a false sense of security and risk compensation.<br>*For example, if interventions to increase pre-exposure prophylaxis (PrEP) for HIV-prevention in men who have sex with men lead to a change in the perception of risk from unprotected sexual intercourse[30-35].* | *"For example, breast cancer prevention messages emphasizing the need for women with a family history of breast cancer to have regular mammography created a false sense of security among women who did not have a family history of breast cancer (Lerman 1991[37]).* |

| Mechanism | Dimension | Description | Example |
|---|---|---|---|
| | Through (self-) labelling, stigmatization and stereotyping | Unintended consequences may arise through the measure affecting or interacting with how individuals or populations are being perceived or are perceiving themselves; in the form of (self-)labelling, stigmatization, or stereotyping. This includes the assignment of responsibility or labels in the form of certain characteristics, as well as the resulting experiences (e.g., guilt and shame).<br>*For example, an obesity-focused public health intervention emphasizing the importance of behaviour change can lead to obese individuals being assigned the responsibility for their body weight (excluding genetic and environmental factors) and as a result the label of other related negative labels such as being lazy, with consequences for their mental health and wellbeing or them being discriminated[16 17 38 39].* | *"In healthcare settings, women who perceive stigmatization from their providers report delaying use of preventive health services for fear of being judged or embarrassed. This avoidance of care allows for untreated problems to progress to a more advanced stage that may be more difficult to treat, thus exacerbating health problems (Stangl 2019[23])* |
| **Through available opportunities** | Definition and description | The range of opportunities to act or react under existing or changing circumstances, which are perceived as available to individuals, populations, or institutions, result from the interaction between the available and accessible resources, the characteristic of the setting (e.g., rules and regulations), and characteristics or knowledge, skills and abilities of the individuals, populations, or institutions. Thus, affecting one of these components can lead to an increase or decrease in the range of opportunities for (re-)action perceived as available to the affected individual, population, or institutions and as a result can lead directly or indirectly (e.g., through reactive behaviour change) to unintended consequences. Furthermore, can unintended consequences arise through changes in the situation or circumstances of individuals, populations, or institutions when an adequate reaction to the change is (perceived as) not possible due to the lack of appropriate available opportunities.<br>*For example, infection control measures such as social distancing or curfews can constrain the option of meeting other individuals. This can lead to the experience of isolation and loneliness as a mental health consequence[40].* | *"Confinement, loss of usual routine, and reduced social and physical contact with others were frequently shown to cause boredom, frustration, and a sense of isolation from the rest of the world, which was distressing to participants. This frustration was exacerbated by not being able to take part in usual day-to-day activities, such as shopping for basic necessities or taking part in social networking activities via the telephone or internet." (Brooks 2020[40])* |
| | Through rules and restrictions and their enforcement | Unintended consequences can arise through the measure affecting or interacting with rules or regulations which define the range of options for behaviours or environments available and accessible to individuals or populations. This includes the enforcement mechanisms in place to ensure adherence, and the consequences resulting from them (e.g., prosecution of individuals not adhering to rules on the consumption of illicit drugs).<br>*For example, the implementation of a curfew as an infection control measure in a pandemic can restrict meetings outside and as a result may lead to more meetings indoors with higher risk of infections.* | *"Covid 19 policy: The United Kingdom and the United States are isolating the elderly and those living in care homes . Evidence of potential harm: Loneliness, depression, need for health and social care, access to care." (Glover 2020[41])* |

| Mechanism | Dimension | Description | Example |
|---|---|---|---|
| | Through knowledge, skills and abilities | Unintended consequences may arise through the measure affecting or interacting with the available opportunities of action resulting from the knowledge, skills or abilities of individuals, populations, or institutions. Measures can affect these through providing (correct or false) information, training or knowledge transfer, or the lack thereof (i.e., loss of ability or experience through lack of training).<br>*For example, a public health campaign leads to reduction in the number of cases of typhoid fever in an area. Due to lack of exposure to typhoid fever cases, the health system, and the stakeholder within them slowly lose their ability to handle these types of cases. Leading to higher rates of death among the (fewer) typhoid fever cases that still occur.* | *"The use of inaccurate information in crisis communications can produce a number of negative consequences. Most important amongst these is a lost opportunity to provide assistance with regards to the crisis at hand and thus prevent harms occurring to individuals and society in general. In addition to the harms that occur to individuals, groups and their interests in society, incorrect communication also represents a waste of public resources that could be used more effectively" (Quinn 2018[42])* |
| | Through available and accessible resources, goods and services | Unintended consequences may arise through the measure affecting or interacting with the availability, accessibility, and affordability of goods or services as well as financial and non-financial resources. This includes co-benefits resulting from increasing availability and access.<br>*For example, an intervention to increase foetal growth in LMICs may increase the risk of obstructed labour, mortality, and morbidity due to the lack of infrastructure to perform caesarean sections in a timely and safe manner[43].* | *"Harsh approaches are not just ineffective; they are often counterproductive. When access to treatment and sterile needles and syringes were restricted in some countries, HIV prevalence increased among people who inject drugs." (Frei 2017[44])* |
| | Through (self-)efficacy and empowerment | Unintended consequences may arise through the measure affecting or interacting with the experience of agency, (self-)efficacy or control over a situation of individuals and populations. This furthermore includes the experience, expectation, or perception of barriers or facilitators (e.g., for barriers for behaviour change), and the perceived ability to overcome them as well as the awareness about precedents, i.e. a case, event or action that is regarded as an example or guide to be considered in subsequent similar circumstances.<br>*For example, after a city introduces new health-related taxes which showed positive financial and health related consequences within this city, other cities close by follow this case study[45].* | *"The unintended effects of the Minnesota Heart Health Program included more than the treatment city of Bloomington's ban of cigarette sales through vending machines. The city's ban provided an impetus for movements to pass similar bans in other cities in the state and for organizing opposition to the efforts of bar, hotel, and restaurant owners to pre-empt local restrictions at the state level". (Cho 2007[45])* |
| **Through environments and environmental exposure** | Definition and description | Unintended consequences may arise when the measure leads to changes of the (natural, physical, or social) environment individuals, populations, or institutions are already exposed to. Furthermore, unintended consequences may arise when individuals, populations, or institutions are more (or less) exposed to environments and environmental risks as a result of the measure. Environmental exposure is defined broadly and includes factors such as exposure to air, atmosphere, chemicals, physical agents, microbiological pathogens, noise, vibration, radiation, temperature, etc. It furthermore includes the exposure to goods and services (e.g., types and quality of food and water), to accidents, or to violence.<br>*For example, providing a financial incentive for physical active mobility to the workplace leads to an increase of individuals cycling to work. Due to an increased exposure to an accident-prone physical activity environment, this can result in an increase in road traffic accidents[10 46].* | *"An increasingly widely held view demonstrates that social, organizational, and physical environments are important determinants of behavior. Environments, and people's perceptions of their environments, may con- strain individuals' behavior even when they are highly motivated. Environment and policy concerns are often central to health disparities: Having access to walkable communities, safe parks, and recreational facilities is associated with more physical activity and lower risk of obesity, but communities of color often have less access to such resources in their neighborhoods." (Glanz 2010[47])* |

| Mechanism | Dimension | Description | Example |
|---|---|---|---|
| | Through changing characteristics of environments | Unintended consequences may arise through the measure affecting or interacting with characteristics of environments. This includes physical changes of environments (e.g., construction and changes of infrastructure) and changes of other natural characteristics (e.g., quality of water).<br>*For example, well digging to increase the access to clean drinking may unintentionally contaminate the groundwater with arsenic or other pollutants (e.g., bacteria, minerals)[48].* | *No suitable paragraph was found. Derived theoretically by discussion within the research team.* |
| | Through changing exposure to environments | Unintended consequences may arise through the measure affecting or interacting with the exposure of individuals and populations to environments. This includes exposure to nature (e.g., UV-light, pollens, weather) and human made emissions (e.g., particulate matter, noise).<br>*For example, a measure to reduce CO2 emissions of cars and associated diseases may also decrease the exposure to noise and other pollutants as well, which may have multiple beneficial health effects.* | *No suitable paragraph was found. Derived theoretically by discussion within the research team.* |
| | Through quality and characteristics of goods or services | Unintended consequences may arise through the measure affecting or interacting with environments and environmental exposure via the quality and characteristics of goods and services.<br>*For example, a taxation of "unhealthy" products to decrease their consumption can lead to the industry reformulating their product to be healthier, including those not affected by the taxation[49].* | *"On the other hand, warnings may also improve health by prompting industry reformulation. Nutrition policies can be designed specifically to encourage product reformulation; doing so may help shift the balance of implied responsibility to an ethically more favorable equilibrium (i.e., toward shared responsibility between individuals and industry). Additionally, warning policies can be accompanied by mass communication campaigns that emphasize that individuals, organizations, industry, and the government each have a role to play in encouraging healthy eating." (Grummon 2020[36])* |
| | Through accidents and violence | Unintended consequences may arise through the measure affecting or interacting with environments and environmental exposure regarding the probability and consequences of accidents and being affected by crime or violence. Measures can affect the probability of accidents or crime occurring, their severity, or the presence and effectiveness of institutions preventing accidents or crime and mitigating their consequences.<br>*For example, public health interventions to increase physical activity can increase the number of individuals cycling, leading to an increase of populations at risk of being affected by road traffic accidents[10].* | *No suitable paragraph was found. Derived theoretically by discussion within the research team.* |
| **Through social norms and practices** | Definition and description | Unintended consequences may arise through the measure affecting or interacting social norms, practices, or relationships. This includes the formation of new and the reformation of existing social norms, roles and identities, as well as social practices arising from them (e.g., | *"Social narratives—such as those which determine the boundaries of 'appropriate' behaviour by pregnant women (eg, around light consumption of alcohol)—may cause stress and guilt even where their putative* |

| Mechanism | Dimension | Description | Example |
|---|---|---|---|
| | | discriminatory practices or institutions). Furthermore, this includes the measures leading to unintended consequences through affecting social networks and relationships.<br>*For example, an anti-smoking campaign to reduce public tobacco smoking can promote changes in social norms and practices. This can result in smokers being perceived as deviant and face social judgement and exclusion[50].* | *health rationales receive limited support from the evidence. Such narratives may also lead people to reject public health messages as irrelevant to them, for example, by perpetuating stereotypes of particular health risks as associated with lower or marginal status." (Lorenc 2015[11])* |
| | Through affecting social roles, norms, and practices | Unintended consequences may arise through the measure affecting social roles and identities, as well as social norms, and the practices resulting from them. Measures can lead to the introduction or establishing new social roles, identities, norms, and practices or through reproducing, reinforcing, or challenging existing ones.<br>This can take place through (i) copying the action of other individuals in an ambiguous social situation where people are unable to determine the appropriate mode of behaviour (social proof), (ii) providing examples for people to aspire to or imitate (modelling), or (iii) through imitating the social practices in a social group and incorporating the norms, values, and social practices associated with them (habit formation).<br>*For example, a public health campaign aimed at mother's who smoke, may establish, or interact with existing traditional social norms of mothers being guardians of family health who are expected to place their children's concerns first. This may lead to the experience of shame or guilt by mothers who smoke or transgressing these social norms. It may furthermore lead to the anticipation of social judgement, resulting in mother's limiting health care seeking behaviours. It may furthermore lead to social judgement and discrimination (e.g., in the form of insult, gossip, or social exclusion) by the society, because of these mothers transgressing social norms[29].* | *"This separation from sexuality education can become a problem when sexual abuse prevention, with vague allusions to private parts and emphasis on saying no, constitutes the first or only classroom reference to sexuality; young children then learn that sexuality is essentially secretive, negative, and even dangerous." (Trudell 1988[51])* |
| | Through social networks and relationships | Unintended consequences may arise through the measure affecting or interacting with social networks and relationships, including the interconnectedness of individuals and populations, as well as the quality of the relationships within them.<br>*For example, infection control measures such as social distancing or curfew regulations during a pandemic may reduce the level of social interaction and the quality of relationships of individuals affected by them, leading to consequences for wellbeing and mental health, such as feelings of boredom, isolation, or depression[40].* | *"Confinement, loss of usual routine, and reduced social and physical contact with others were frequently shown to cause boredom, frustration, and a sense of isolation from the rest of the world, which was distressing to participants." (Brooks 2020[40])* |
| | Through discriminatory practices or institutions | Unintended consequences may arise through the measure affecting or interacting with discriminatory social practices, including "enacted stigma" as discriminatory practices or sanctions on individual and collective levels. Consequences may arise from individuals or populations acting towards others based on bias or on stigma, including self-disabling and self-harming behaviours based on internalized stigma. | *"Beyond the modification of medical office equipment to meet the needs of larger patients, both the stigmatizing effect of current medical language and the pathologization of 'fatness' may lead to strong negative responses by patients. Stigmatized care provider attitudes are picked up by* |

| Mechanism | Dimension | Description | Example |
|---|---|---|---|
| | | *For example, a public health measure may enforce discriminatory attitudes in a society against smokers. The resulting discriminatory practices may lead to affected individuals feeling social isolation or adverse mental health consequences.* | *patients, and in the case of children, by their parents." (MacLean 2009[52])* |
| **Through economic mechanism** | Definition and description | Unintended consequences may arise through the measure affecting (i.e., creating or restricting) or interacting with economic mechanisms and processes. This includes incentives or disincentives as well as price and market mechanisms resulting from the balance between the balance and supply of goods and services.<br>*For example, a public health program which provides a bounty for a killed cobra to reduce the risk of snake bites, may lead to an increase due to the population incentivised to engage in the breeding of cobras (these are so called perverse incentives).* | *"Food warning policies could also spur manufacturers to reformulate products to avoid triggering a mandatory warning.42,43 While research on food warnings' effects on reformulation is limited, studies examining other nutrition labeling schemes, as well as other nutrition policies like taxes, find that implementing these policies can prompt positive changes in products' overall nutritional profile. " (Grummon 2020[36])* |
| | Through incentives or disincentives | Unintended consequences may arise through the measure affecting or introducing incentives or disincentives. In this context, incentives refer to the expectation of financial or economic gain or rewards (including social capital) and disincentives to the expectation in cost or punishment.<br>*For example, a public health program which provides a bounty for a killed cobra may not lead to a decline in the cobra population, but to an increase due to the population incentivised to engage in the breeding of cobras (these are so called perverse incentives).* | "In Australia, expenditure on tobacco has a significant negative association with expenditure on nearly all household items (including necessities such as clothing, footwear, and food). In New Zealand it has been estimated that tobacco spending involved up to 14% of the non-housing budgets of households with smokers (i.e., for those in the second lowest income decile). These findings indicate that tobacco tax is contributing to financial hardship among low-income households in some settings. (Wilson 2005[8]) |
| | Through demand, supply, and their balance in markets | Unintended consequences may arise through the measure affecting the need and demand or the supply and capacity of resources, goods, or services, as well as the balance between these two (e.g., in the form of increasing or decreasing prices).<br>*For example, warning labels on sugar-sweetened beverages leading to a reduction in demand. In response, the prices of the products may decrease, providing incentives of an (re-)increase in the consumption of these beverages.* | *"Studies of screening programs found such interventions can increase the burden or perceived burden on referral systems by causing increases in referrals beyond the capacity of the system when adequate procedures have not been put in place for handling referral increases due to positive screens." (Kuiper 2018[53])* |
| **Through the functioning of systems** | Definition and description | Unintended consequences may arise through the measure affecting or interacting with the functioning of systems (e.g., health system), including single sub-systems of bigger systems (e.g., primary schools within the educational systems; or insect populations within an ecological system). In this context, systems can refer to social, economic, political, organizational, or ecological systems.<br>This includes the resilience, resistance, or sustainability of systems, the creation of synergies or antagonistic effects across systems, as well as the functioning of systems and its components. | |

| Mechanism | Dimension | Description | Example |
|---|---|---|---|
| | | *For example, a syringe exchange program can serve as a delivery platform for other interventions or services (e.g., vaccination services). This can result in an increased utilisation of other interventions or health care services.* | |
| | Through affecting the functioning of system component and subsystems | Unintended consequences may arise through the measure affecting or interacting with the functioning of systems, including single sub-systems of a bigger sytems (e.g., primary schools within the educational sytems; or insect populations within an ecological system).<br>Unintended consequences may arise from a measure affecting a component of a system, leading to consequences beyond the direct effect of the measure or through affecting the functioning of the system or other system components. If an agent, resource, practice, or institution, which serves a particular function in an individual or system, is affected (e.g., removed) this can disrupt the functioning of the system as a whole; leading to additional unintended consequences. Unintended consequences may further arise through the measure affecting or interacting with sub-systems of a bigger systems (e.g., insect populations within an ecological system). This includes human made systems (e.g., health systems) as well as ecologic systems.<br>*For example, US-policies to reduce wildfires led to fewer fires, but also led to growth conditions and a build-up of dead wood, resulting in a condition in which when wildfires occurred, these were much larger and more devastating. The small fires not only served a function in the ecosystem, but they also prevented larger fires[54].* | *No suitable paragraph was found. Derived theoretically by discussion within the research team.* |
| | Through resilience, resistance, or sustainability of systems | Unintended consequences may arise through the measure affecting or interacting with the resilience, resistance, or sustainability of systems. While the system may not be affected under "normal" circumstances, stressors on the system may lead to collapse and affect the ability to recover.<br>*For example, if a health care reform reduces the availability of ICU beds in a health system as a cost saving measure without adversely affecting the treatment of patients under normal caseloads. This reduced capacity can increase the risk of the health care system collapsing under the stress of an extreme event, such as a pandemic, increasing the morbidity and mortality burden.* | *No suitable paragraph was found. Derived theoretically by discussion within the research team.* |
| | Through creating synergies or antagonistic effects | Unintended consequences may arise through the measure interacting with another public health measure or system component, leading to a synergistic or an antagonistic interaction (or the lack thereof).<br>*For example, a specific public health intervention (e.g., syringe exchange programs) can be used as a delivery platform for other interventions or services (e.g., vaccination services).* | *„For example, syringe-exchange programmes for injection drug users can serve as delivery sites for other health services, and help gain the trust of this under-served population.“ (Guttmann 2004[5])* |

### Supplementary File 8: Application of CONSEQUENT

#### Supplement 8.a: Simplified guidance on how to apply the framework in a conceptual manner

(1) Start by developing a process-oriented logic model[9 55] or a complex systems map of how the intervention is meant to work within the context it is implemented in. Logic models represent graphical depictions describing the intended impacts of the interventions, along with the assumed causal pathway(s). The proposed conceptual application of the framework builds on the suggestions made by many authors to expand the logic models to also include plausible unintended consequences and the associated mechanisms. To assist users, we proposed the following steps:

(2) Expand the model by theorizing which unintended consequences could arise from the intervention. The proposed CONSEQUENT framework is intended to support this process in two ways. First, the framework component on the consequences provides a list of potential unintended consequences to be used in theorising (e.g., what unintended ecological consequences could arise from travel restrictions as an infection control measure?[56]). Second, the framework component on mechanisms provides a list of mechanisms to support reflecting on the processes that could be triggered by the introduction of the measure into the system and the potential consequences as well (e.g., could rapid testing as an infection control measure in a pandemic affect the perception of risk and because of risk compensation behaviour change to an increase in the number of infections?[57]).

(3) Conduct a mapping exercise to identify (sub)populations which are or could be affected by the PH intervention in a particular manner. In identifying these populations, frameworks, ss PROGRESS+[58] can work as helpful tools (see, for example, Glover et al.[59]). Revisit the logic model keeping the individual (sub)populations in mind and adapt or expand the logic model accordingly (e.g., are there specific unintended consequences of school closures as a pandemic control measure for individuals with a low socio-economic status?)

(4) Examine publications on similar measures, in particular regarding the observed unintended consequences. The required degree of similarity and directness of the evidence on the measure of interest is likely to vary (e.g., whether the evidence on stigmatization resulting from media campaigns on HIV can contribute insights on unintended consequences regarding the media campaigns on obesity). Revise and expand the logic model accordingly.

(5) Integrate e the perspectives of affected stakeholder groups, including individuals or groups with particular insights into the specific contexts and how interventions might operate within them[9], as well as the views of those opposing the intervention[60].

While presented linearly, we assume that engaging in the process in an iterative manner and repeating different steps (e.g., revisit the framework after or in consultation with affected stakeholders) will improve the final logic model.

#### Supplement 8.b: Exemplary application of the CONSEQUENT framework

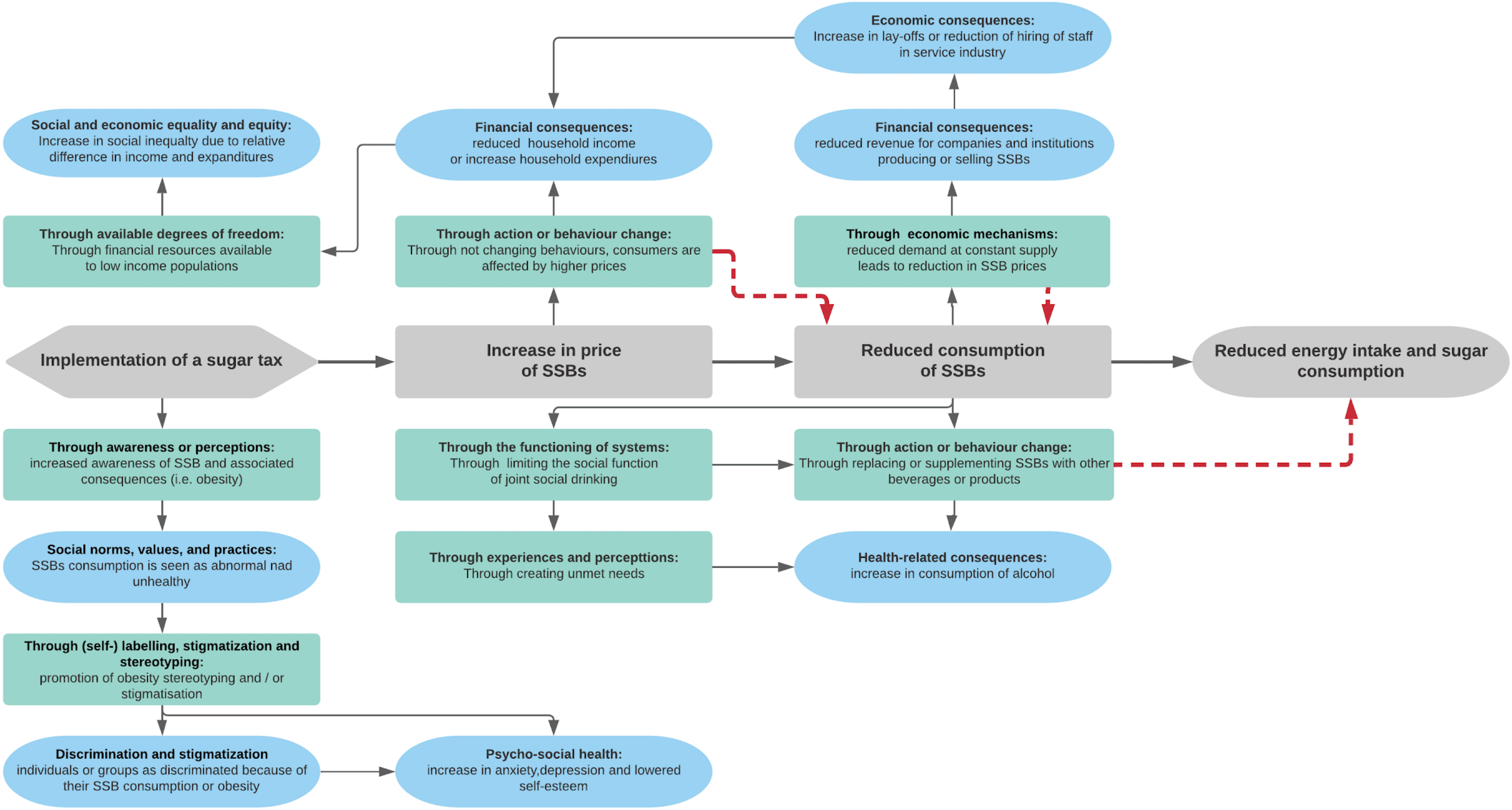

Figure A4.1: Simple example of a logic model of unintended consequences to depict the conceptual use of the framework
